# The Evolutionary History of Metastatic Pancreatic Neuroendocrine Tumours Reveals a Therapy Driven Route to High-Grade Transformation

**DOI:** 10.1101/2024.01.08.24300723

**Authors:** Samuel Backman, Johan Botling, Helena Nord, Suman Ghosal, Peter Stålberg, C. Christofer Juhlin, Jonas Almlöf, Anders Sundin, Liang Zhang, Lotte Moens, Barbro Eriksson, Staffan Welin, Per Hellman, Britt Skogseid, Karel Pacak, Kazhan Mollazadegan, Tobias Åkerström, Joakim Crona

**Author notes:** **Corresponding author:** Joakim Crona, Akademiska Sjukhuset ing 100 75185 Uppsala, Sweden. **Conflicts of interest:** The authors have no conflicts of interest to declare.

## Abstract

Tumour evolution with acquisition of more aggressive disease characteristics is a hallmark of disseminated cancer. Metastatic pancreatic neuroendocrine tumours (PanNETs) in particular, show frequent progression from a low/intermediate to a high-grade disease. To understand the molecular mechanisms underlying this phenomenon, we performed multi-omics analysis of 32 longitudinal samples from six metastatic PanNET patients. Following MEN1 inactivation, PanNETs exhibit genetic heterogeneity on both spatial and temporal dimensions with parallel and convergent tumuor evolution involving the *ATRX/DAXX* and mTOR pathways. Following alkylating chemotherapy treatment, some PanNETs develop mismatch repair deficiency and acquire a hypermutator phenotype. This DNA hypermutation phenotype was only found in cases that also showed transformation into a high-grade PanNET. Overall, our findings contribute to broaden the understanding of metastatic PanNET, and suggests that therapy driven disease evolution is an important hallmark of this disease.

## Main

Pancreatic neuroendocrine tumours (PanNETs) patients display extraordinary diverse disease characteristics^1,2^: This neoplasm may secrete various neuropeptides and demonstrate a wide spectrum of invasiveness and cell proliferation that ranges from slowly growing localized lesions, to rapidly dividing disseminated tumours^3,4^. These factors are recognized as prognostic markers determining the outcome from the disease. Further, many PanNET patients experience evolving tumour characteristics over time with changes in hormone secretion pattern and/or tumour grade. This evolutionary phenomenon is usually associated with a very poor prognosis^4–7^. The underlying mechanisms leading to such changes in PanNET biology are not understood.

The genetics of primary PanNET is well researched with mutations in *MEN1* as well as genes related to mechanistic target of the rapamycin (mTOR) signalling pathway, and chromatin remodelling (*ATRX/DAXX*) as major hallmarks^8,9^. Somatic inactivation of *ATRX* and *DAXX,* with subsequent activation of the alternative lengthening of telomeres phenotype, is suggested to occur as secondary events in PanNETs^10–12^. It was also shown that primary PanNETs with loss of DAXX, ATRX, H3 lysine 36 trimethylation, ARID1A, and/or CDKN2A are associated with shorter survival^13^. Pan-genomic data on metastatic PanNET is still lacking and disease evolution has so far only been studied using targeted DNA sequencing: While most PanNET appears to have stable genomic landscapes over time, some tumours may be influenced by chemotherapy-associated mutagenesis^13–15^. As such, more information is clearly needed to understand why PanNET may transform and acquire more aggressive traits. Hence, we performed a detailed pan-molecular investigation of multiregional PanNET samples that had been collected over the course of the disease, before and after treatment with systemic therapies.

## Results

To characterise the biology and evolution of metastatic and lethal PanNET we performed multi-omics analysis of 32 primary and metastatic tumour samples representing 18 unique lesions from 6 patients (Figure 1A, Supplementary Figure 1, Supplementary Tables 1-2). All patients had metachronous samples with 19 collected at baseline and 13 at follow-up.

**Figure 1.**
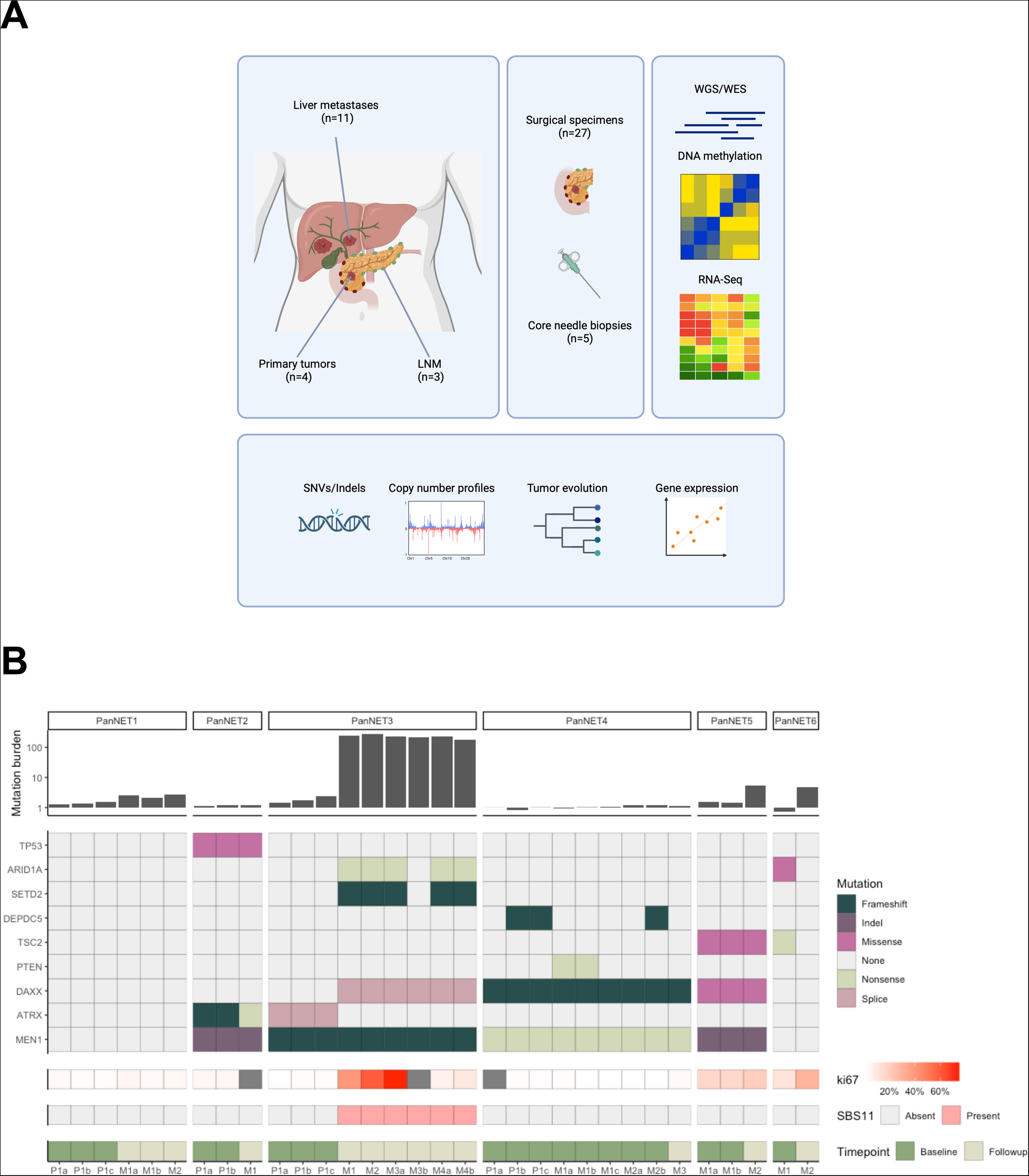
a) Overview of experimental strategy. Created with BioRender.com. b) Summary of main findings across the cohort. PanNET samples were annoteted for location (P, Primary tumour; M, metastasis), Tumour lesion number (1-4) and Sample number from each individual tumour lesion (a-c). Abbreviations: LNM, lymph node metastasis; WES, Whole exome sequencing; and WGS, Whole genome sequencing.

Tumours were WHO grade 1-2 at diagnosis and three patients had hormonal syndromes. None fulfilled criteria for inherited tumour syndromes and all were negative for cancer-associated pathogenic germline pathogenic variants (Supplementary materials). While two patients had been exposed to systemic therapies before acquisition of baseline tumour samples, all had been exposed to various PanNET approved systemic therapy regimens before collection of follow-up tumour samples.

### Genetic landscape and evolution in metastatic PanNET

Whole genome (*n*=27, median 67X) and whole exome (*n*=5, median 438X) DNA sequencing revealed a genetic landscape representative of PanNET with pathogenic variants in established disease drivers in all but one patient (Figure 1B, Supplementary Figures 2A-F, Supplementary Tables 3 & 4). These results align with previous studies with four out of six patients carrying somatic *MEN1* mutations, while somatic pathogenic variants in *ATRX*, *DAXX* and the mTOR pathway were common auxiliary events. *MEN1* alterations were fully clonal in 4/4 patients with remaining pathogenic variants in PanNET genes occuring in a subclonal fashion: *ATRX*/*DAXX* 2/4 fully clonal, mTOR related genes 1/3 fully clonal and chromatin integrity related genes (*ARID1A*/*SETD2*) 0/2 fully clonal (Supplementary Figures 3A-D). To further describe the evolutionary histories of PanNETs, we used these high confidence substitutions and insertion-deletions to derive phylogenies, which showed complex and heterogenous architectures: A monoclonal seeding pattern could be confirmed in two patients that both had one distinct clone present in all metastases (Figures 2C and 3). Contrary, PanNET04 displayed a polyclonal seeding pattern with two distinct metastatic clones originating from separate parts of the primary tumour (Figure 3C). Here the two clones showed evidence of convergent evolution on the mTOR pathway with parallel independent truncating mutations in *DEPDC5* and *PTEN.* The tree topology of PanNET04 showed two distinct branches, each containing both samples from the primary tumour and from metastatic lesions. Such a branched evolution pattern could also be observed at the *ATRX*/*DAXX* loci: for PanNET02 with different *ATRX* pathogenic variants in primary tumour and metastasis as well as for PanNET03 with *ATRX* mutated primary tumour samples and a distinct *DAXX* mutation observed in all samples from its metastases (Figures 2C and 3B). For two patients, whole exome sequencing data was available from metastatic lesions: In PanNET05 all samples were found to share pathogenic variants in *MEN1*, *DAXX* and *TSC2* while the two metastases from PanNET06 had discordant mutation status for mutations in *TSC2* and *ARID1A*, which were found in the baseline sample only (Figure 1B, Supplementary Figures 3E-F).

**Figure 2.**
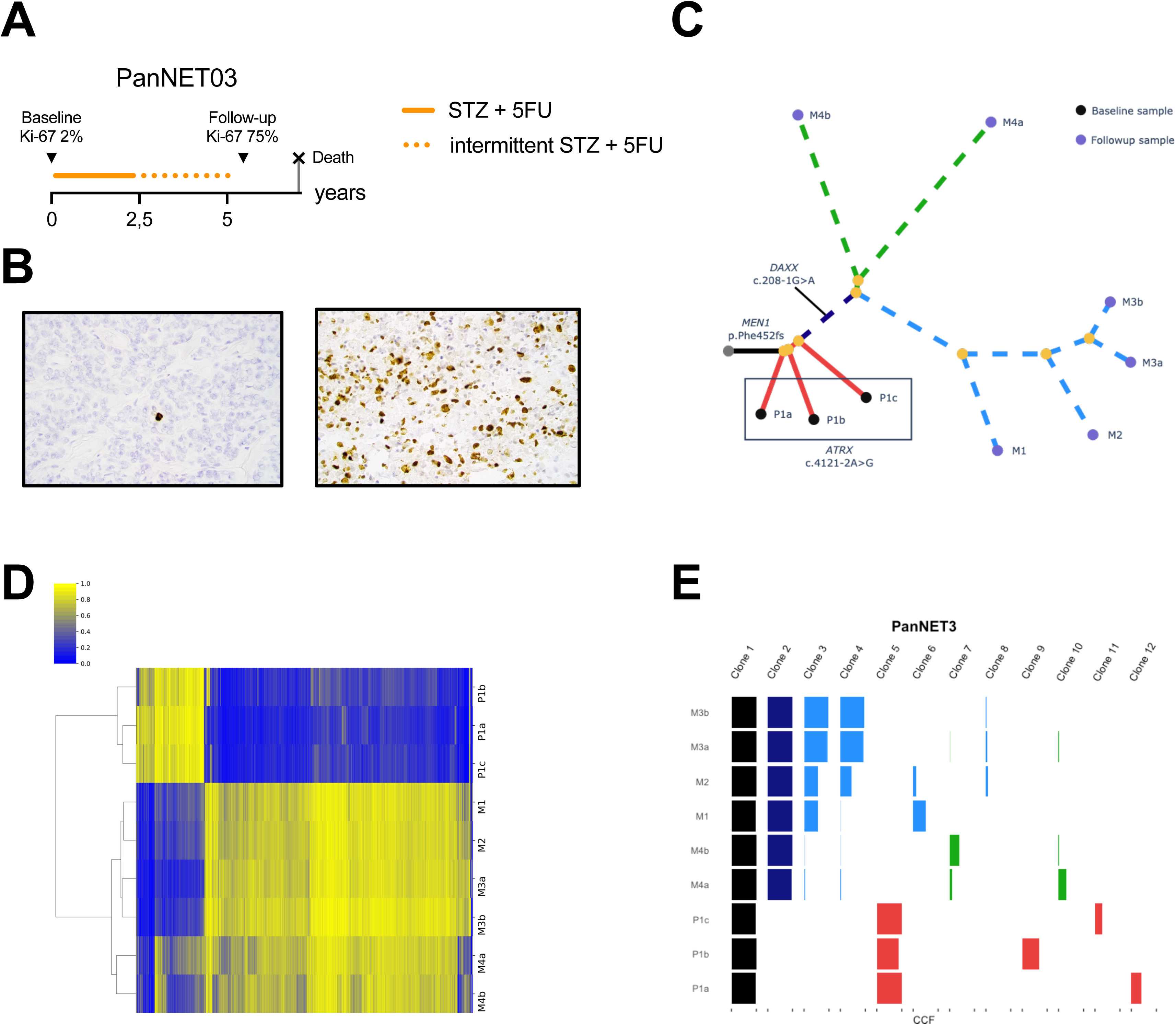
a) Treatment timeline for patient PanNET03. b) Representative images of Ki67 staining of primary and metastatic samples P1a and M3a demonstrates high-grade transformation with Ki67 rising from 1% to 75%. c) Phylogenetic tree of bulk samples from PanNET03. PanNET samples were annoteted for location (P, Primary tumour; M, metastasis), Tumour lesion number (1-4) and Sample number from each individual tumour lesion (a-c). The metachronous metastatic samples form an outgroup and convergent evolution of the *ATRX/DAXX* is seen. The dotted lines have been shortened for visualization purposes. d) The metastatic samples share a distinct DNA methylation pattern. e) Subclonal reconstruction with PyClone-VI reveals separate dominant clones shared by the primary tumour and metastatic lesion samples respectively. Abbreviations: CCF, Cancer cell fraction; and STZ, streptozocin.

**Figure 3.**
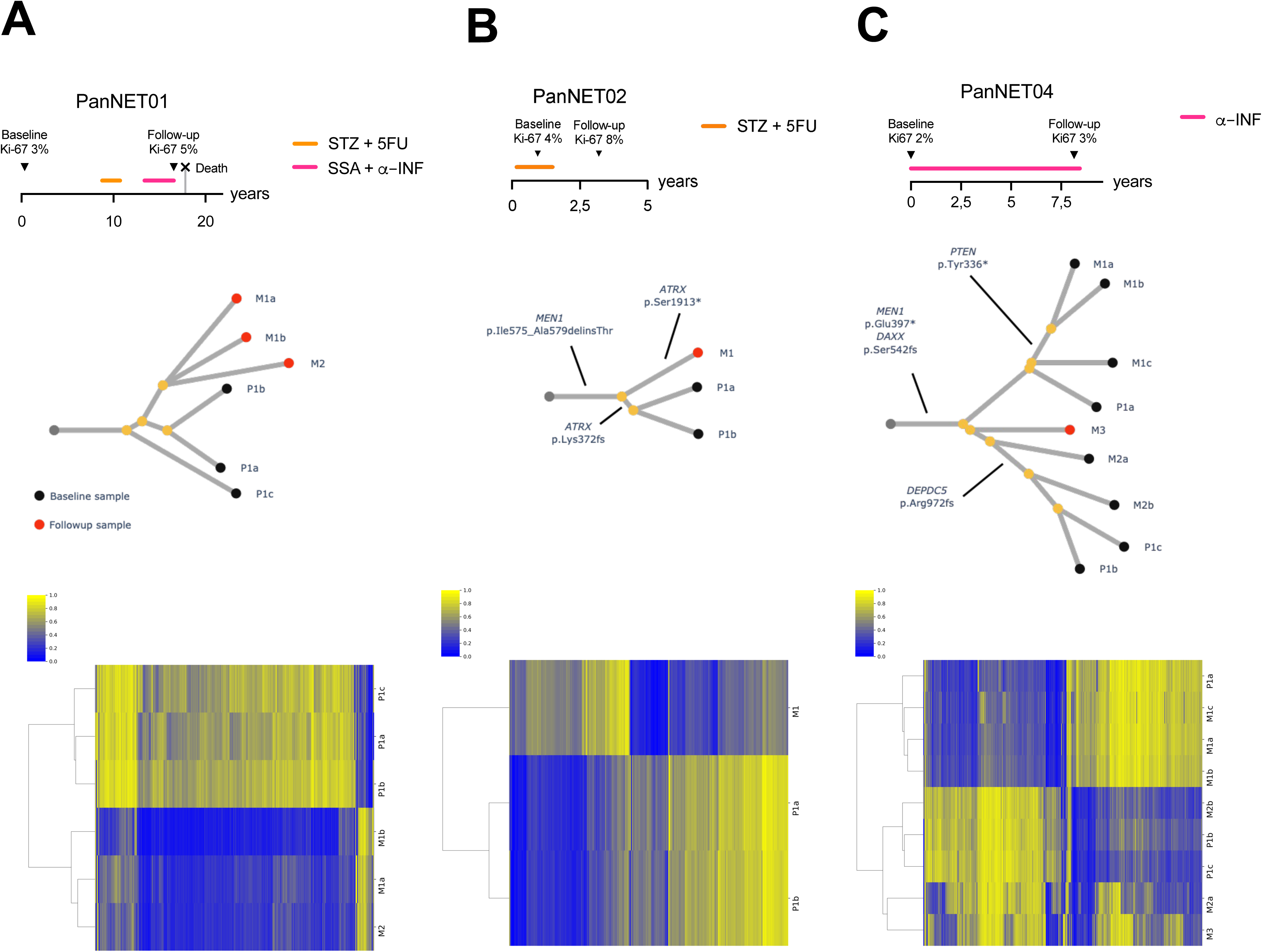
a-c) Timelines, phylogenetic trees, and DNA methylation heatmaps for PanNET01, PanNET02 and PanNET04. PanNET samples were annoteted for location (P, Primary tumour; M, metastasis), Tumour lesion number (1-4) and Sample number from each individual tumour lesion (a-c). Groupings based on DNA methylation and on somatic mutations are noted to show the same patterns. For PanNET04, the lengths of some branches have been increased for visualization purposes. The corresponding raw trees are available in the supplementary material. Abbreviations: α-INF: Alfa-interferon; SSA, somatostatin analogue; STZ, streptozocin.

The tree topology based on small mutation data was largely concordant with the tree topology resulting from DNA methylation data (Supplementary Table 4): In all cases we found a striking similarity between the two reconstructions and clades of closely related samples clustered together in the DNA methylation analyses (Figure 2D and Supplementary Figures 4A-H). On the cohort level, all tumour samples clustered together with other samples from the same patient based on both DNA methylation and gene expression data (Supplementary figures 5 and 6C).

In order to further characterize the evolution of metastatic PanNET we performed computational inference of subclones using PyClone-VI that recapitulated the results from the sample-level phylogenetic analysis (Figure 2E, Supplementary figures 7A-C). In PanNET01, the primary tumour samples shared a subclonal cluster of mutations, believed to give rise to the metastatic samples. The metastatic samples shared additional mutations not present in the primary tumour. In PanNET02 and 03, the primary tumour samples shared a clonal cluster not present in those from metastases, suggesting early metastatic seeding which predates the most recent common ancestor of the primary tumours (Figres 2E and 3B). In PanNET04, parallel metastatic seeding from different geographical regions of the primary tumour was observed, with one of the primary tumour samples (Sample P1a) sharing a cluster with three samples from a liver metastasis, while two other samples from the primary tumour shared mutation clusters with a retroperitoneal lymph node metastasis (Figure 3C). A second liver metastasis did not harbour either of these mutation clusters, suggesting seeding from a separate non-sequenced part of the primary tumour, or prior to the establishment of the clones dominating in the primary tumours.

Our results confirm that *MEN1* inactivation occurs very early in tumourigenesis, with *ATRX/DAXX* being inactivated in subsequent steps. We then sought to determine the timing of the conserved choreography of chromosomal mis-segregations previously described in PanNETs, that has been linked to inactivated *ATRX/DAXX* leading to chromosomal cohesion failure^16^. Patients PanNET03 and 04 had this chromosomal mis-segregation pattern, and by performing binomial modelling based on tumour purity and ploidy as well as allele frequency data we show that *ATRX*/*DAXX* mutations occurred before the whole genome doubling event leading to copy number neutral loss of heterozygosity (Supplementary results). Moreover, we were able to confirm that the major loss of heterozygosity events affect the same physical chromosome across samples of the same patient (Supplementary figure 15). This is consistent with one catastrophic event during tumour evolution, rather than multiple separate events converging on the same copy number profile. These findings corroborate the evolutionary model of PanNET where an islet cell inactivates *MEN1* as *the* tumour initiating event, and that the tumour during its evolution may take different pathways. Inactivation of *ATRX*/*DAXX* is commonly seen and followed by a complex pattern of chromosomal alterations that are associated with aggressive tumour biology^11^. Finally, alterations associated with the mTOR signalling pathway and chromatin regulation often occur late in tumour development, similar to observations across other cancers^17^.

### Treatment induced DNA hypermutation

There was a clear trend towards increased tumour mutation burden (TMB) in metastases compared to primary tumour samples. This was also noted in metachronous samples that had been exposed to different PanNET approved therapies (Figure 1B): Five patients had been treated in accordance with ENETS guidelines^18^ with the alkylating agents streptozocin or temozolomide, and one (PanNET03) developed a hypermutation phenotype with a >100-fold increase in TMB in the follow-up, compared to baseline samples. In order to examine the mechanisms leading to this hypermutated state, we extracted *de novo* mutational signatures from whole genome sequencing mutation sets. Here we identified that a majority of calls in the hypermutated samples could be attributed to a mutation signature resembling SBS11, which has been attributed to treatment with alkylating agents (Figure 4A-B, Supplementary Figures 8A-G), and is characterized by C>T transversions. This signature was largely absent from the non-hypermutated samples, whose mutations were attributed to a flat signature. This finding was corroborated by fitting the mutation sets to COSMIC version 3 signatures (Supplementary Figures 8H-I), the hypermutated samples from PanNET03 showed a majority of mutations attributed to SBS11. The findings in the remaining samples overlapped with primary PanNETs showing SBS 1, 5 and 8 ^8,19^. This SBS11 signature was not present in baseline samples of patient PanNET03, that had not been subjected to streptozocin treatment.

**Figure 4.**
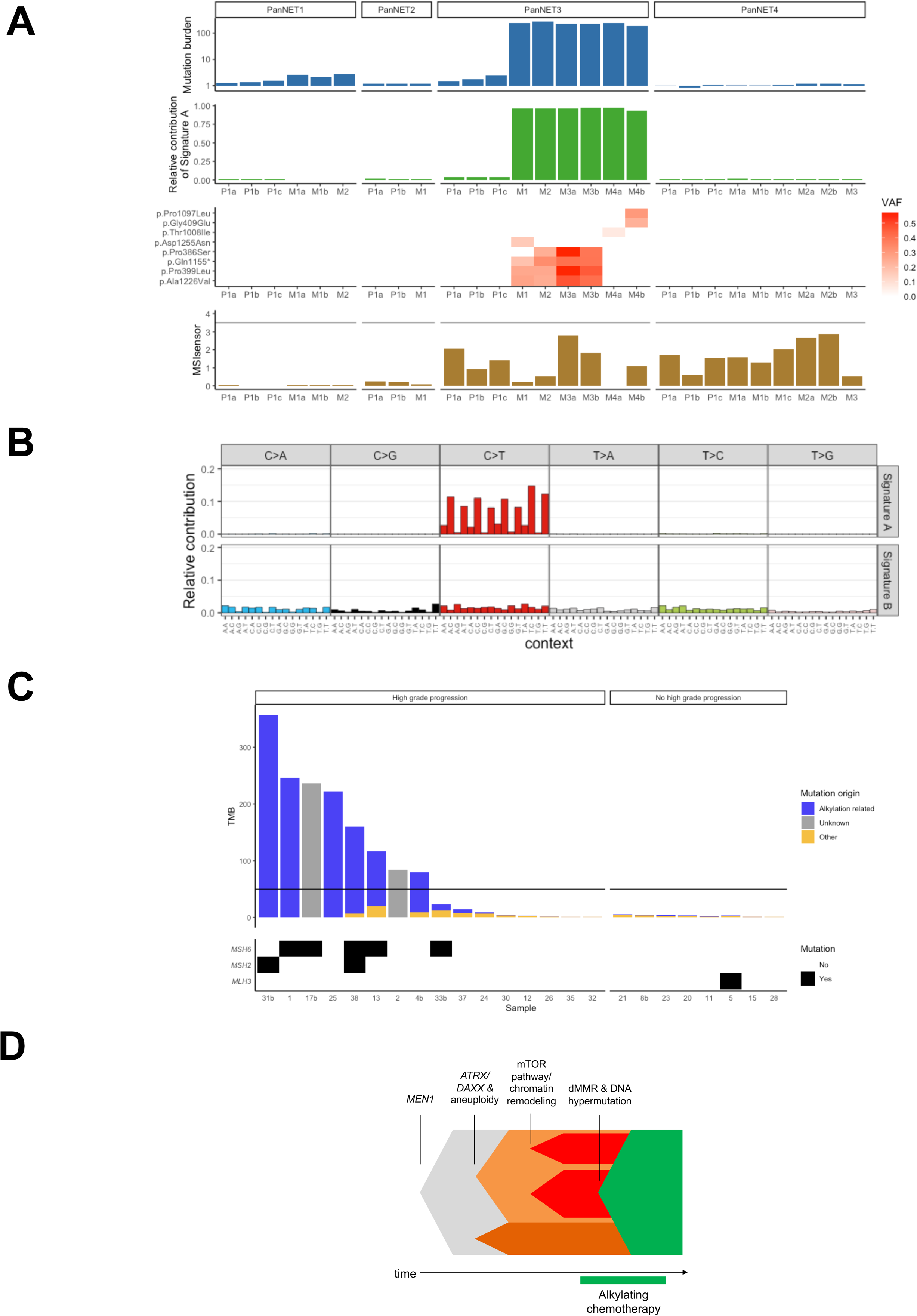
a) Metastatic tumours from PanNET03 exhibit a hypermutation phenotype attributable to a unique mutation signature. PanNET samples were annoteted for location (P, Primary tumour; M, metastasis), Tumour lesion number (1-4) and Sample number from each individual tumour lesion (a-c). The presence of this signature correlates strongly with the presence of pathogenic variants in *MSH6* and is unrelated to microsatellite instability as measured by MSIsensor b) De novo mutation signature extraction of genome sequencing samples identifies a signature dominated by C>T transversions as well as a “flat” signature. c) Tumour mutation burden, alklylation-realted mutation signature and mutation status of mismatch repair genes among PanNET with and without high-grade progression. D) Proposed model for PanNET evolutionary history including the influence of alkylating chemotherapy. Abbreviations: dMMR, mismatch repair deficiency; TMB, Tumour mutation burden; VAF, Variant allele frequency.

We therefore hypothesized that the SBS11-related hypermutation observed in tumour samples of PanNET03 occurred secondary to treatment with streptozocin in the context of mismatch repair deficiency (dMMR), as has been shown in glioblastoma^20,21^.

In order to test the hypothesis that hypermutation is related to dMMR, we scrutinized the hypermutated samples for evidence of damaging mutations in mismatch repair genes (Figure 4A, Supplementary table 5). Samples from tumours M1-M3 shared several mutations in *MSH6*, including a p.Gln1155* nonsense mutation. The samples from the metastatic lesion M4 carried different pathogenic variants in *MSH6*: M4a carried a p.Thr1008Ile mutation previously described as recurrent in hypermutated gliomas^20^, while M4b harboured p.Gly409Glu and p.Pro1097Leu mutations. The finding of different dMMR mechanisms in metastatic samples suggests that the hypermutation phenotype arose comparatively late in the evolution of the tumour, and on separate occasions. To investigate this further, we examined private and shared mutations from the metastases separately, and found that the mutations shared by all metastatic lesions were best fitted to the flat signature seen in the primary tumour samples (Supplementary figures 9A-B). Conversely, mutations common only to M1-M3, as well as those unique to the two samples from M4, could largely be attributed to the SBS11-like signature.

We further analysed whether the hypermutation signature could be attributed to *MGMT* inactivation by promoter methylation, as MGMT is a key enzyme in the repair of alkylated DNA and a mediator of resistance to alkylating agents^22^. This promoter region had variable methylation levels across the cohort (Supplementary Table 6). There was no apparent relation to hypermutation, suggesting that it was not the causative mechanism. In line with previous observations in glioma^20^, no microsatellite instability could be seen in the bulk sequencing data from the hypermutated samples (Supplementary Table 4).

We point to the association between this extraordinary genomic drift fueled by alkylating agents and a large change in clinical behaviour of PanNET03, in whom the disease showed fast progression and high-grade transformation leading to short patient survival. This transformation in tumour biology was confirmed by the increase in Ki67 index from a median 2,2% in primary tumour samples at baseline to a median 40% in follow-up metastatic samples on PanNET03 (Figure 2A). The differences in biology between follow-up samples were also captured on the DNA methylation level: follow-up samples formed a distinct group using unsupervised hierarchial clustering (Figure 2D and Supplementary Figures 4E and F). These differences were most pronounced in regions outside CpG Islands (mean difference 0.13 vs 0.04, p<2.2*10^-16^ by t-test, Supplementary Figure 10). Finally, we could identify differences in gene expression in follow-up samples compared to baseline samples (Supplementary Figure 6A-C). This involved differences in pathways associated with cell proliferation and mesenchymal to epithelial transition (Supplementary table 7), as well as neuroendocrine differentiation and tissue remodelling (Supplementary table 8).

While any direct correlation to genomic variants is nearly impossible in this hypermutated context, the genomic drift was clearly associated with very different biological capabilities. We hypothesized that these features would change the immune landscape, and that PanNET could become susceptible to immune checkpoint inhibitors that are approved across TMB high metastatic cancers. We performed clustering of immune cell infiltrations using CIBERSORT across PanNET samples and identified 3-4 clusters (Supplementary Figures 11A-B and 12). This revealed a heterogenous immune landscape in PanNET03 with hypermutated metastatic samples generally immune depleted, both in the context of pan-cancer samples as well as in our PanNET multi-omis cohort. Analysis with ESTIMATE algorithm for tumour immune infiltration score and stromal score, showed that cluster 1 had a high fraction of immune infiltration, and cluster 2 comprised a high fraction of stromal cells, while cluster 3 demonstrated the highest purity (low immune score, low stromal score and low ESTIMATE score). Again, this feature is in line with that of hypermutated glioblastomas^20^.

### Mismatch repair deficiency, DNA hypermutation and high-grade proression in metastatic PanNET treated with alkylating chemotherapy

To validate the impact of alkylating chemotherapy associated dMMR, DNA hypermutation and high-grade progression in PanNET, we investigated 29 post akylating therapy samples from 24 PanNETs patients (validation cohort 1). Tumour mutation burden was ≥50 in 8/24 (33%) patients, five of whom (63%) harboured mutations in *MSH6* or *MSH2* (Supplementary Table 9). Five patients had two longitudinal follow-up PanNET samples characterized: TMB ≥50 could be detected in three patients. All displayed distinct heterogeneity for both dMMR and DNA hypermutation, which could only be seen in the second follow-up sample. All eight patients with PanNET TMB ≥50 had progression from a low/intermediate to a high-grade tumour (Figure 4C, Supplementary Table 9).

In total, the validation cohort included 16 PanNET patients with high-grade progression, 8 of which had TMB≥50 (50%). Among the eight patients that did not show high-grade progression, none had TMB≥50 (0%, (Odds ratio “infinite” (95% confidence interval 1.8-“infinite” *p*=0.02).

Among primary PanNET samples with DNA sequencing data in the American Association for Cancer Research Project GENIE repository there were only 6/599 (1%) with a TMB of ≥50, all but one of these had an *MSH6* variant (Supplementary Figures 13A-D)^23–25^. Finally, we were able to recover PanNET tissue from one patient with Lynch syndrome due to a pathogenic germline *MSH6* variant: Whole genome sequencing revealed a TMB of 1 and no trace of signature SBS11. This corroborates that SBS11 hypermutation is unlikely to occur spontaneously in PanNET (Supplementary Figure 14). This is in line with previous results in glioma where SBS11 primarily appears in MMR-deficient post-treatment samples^20^.

Together, these experiments allow us to propose the most detailed model of PanNET evolution (Figure 4D) to date. This opens a new field in PanNET research with therapy driven evolution emerging as an disease hallmark with potential detrimental consequences to patients.

## Discussion

Metastatic PanNET is a lethal disease that frequently becomes more aggressive over time. Our work highlights that metastatic PanNET is also a very dynamic disease on the genetic level, with a clear association between genetic drift and more aggressive disease characteristics. This insight broadens our understanding of this lethal disease, with direct implications for biomarker driven research on PanNET. Further, our data indicate how systemic therapy may lead to potential negative consequences for PanNET patients, with alkylating chemotherapy resulting in acquired dMMR, DNA hypermutation and high grade progression.

### Limitations

This is an hypothesis generating study and has its limitations: The small sample size and selection bias of the discovery cohort impedes the detection of novel metastasis-associated events and evolutionary mechanisms. This limitation can largely be attributed to the rarity of cases fulfilling the inclusion criteria in retrospective cohorts. Potential biases due to our limited sample size are accentuated by the mixed nature of the cohort which includes both formalin-fixed and paraffin-embedded core needle biopsies, as well as fresh frozen surgical specimens. The core needle specimens were typically of lower tumour purity, which was partly rectified by the higher depth provided by exome rather than genome sequencing.

Futher, generalizability of our results is limited by the single center design of our study with a potential for unknown center specific practises shaping tumour evolution. Future multicentre prospective studies where tumour tissue is collected according to predefined protocols, including autopsy may help further characterize the evolution of metastatic PanNETs.

Nevertheless, we made an attempt to mitigate these limitations by confirming our key finding of therapy-associated DNA hypermutation in both published datasets and in a set of additional metastatic PanNET samples.

### Interpretation

Our data supports the PanNET evolutionary model where an islet cell undergoes inactivation of *MEN1* as *the* tumour initiating event. The tumor may then take different evolutionary trajectories, which may include inactivation of *ATRX*/*DAXX* followed by a complex pattern of chromosomal alterations, that has associated with a more aggressive tumour biology^11^. Finally alterations involved in the mTOR signalling pathway and with chromatin regulation often occur late in PanNET development, similar to observations across other cancers^17^. However, the most critical finding of this study is how therapy driven tumour evolution may lead to negative consequences for PanNET patients, with alkylating chemotherapy beeing associated with acquired dMMR, DNA hypermutation and high-grade progression. While this finding is strongly supported by unpublished data on PanNETs^26^ it is a well-documented phenomenon in gliomas^20^. Here the selective pressure exerted by alkylating chemotherapy selects dMMR clones, and hypermutation is induced by the dual presence of dMMR and alkylating agents^20^. This is likely due to the inability of dMMR cells to repair the damage incurred by DNA alkylation. As found in glioma, our study suggests that these mechanisms leads to a more aggressive PanNET. Although, future studies is needed to confirm these findings.

Another important question is whether treatment-related hypermutation renders the metastases susceptible to immune checkpoint inhibitors. Although limited data suggests that this is not the case in glioma^20^, experimental evidence implies that the lack of immune activation in hypermutated cases is due to the central nervous system microenvironment^27^. While increased intratumoural heterogeneity is likely to reduce tumour immunogenicity^28^, an initial analysis of the ARETHUSA trial, in which patients with mismatch repair-proficient colorectal carcinoma are treated with temozolomide to induce hypermutation, followed by pembrolizumab, suggests that a subset of patients may achieve disease stabilization with this strategy^21^. Evaluation of immunotherapy in the context of PanNETs with treatment-induced hypermutation may thus be warranted^29,30^. Another hypothesis generated by our data is that slowly proliferating PanNET should not be prioritized for DNA damaging treatments that may accelerate tumour evolution towards more aggressive disease characteristics. Instead patients may have better long-term outcome if surgery and/or somatostatin analogue based therapies are prioritized.

Our results also contextualizes how on-going research efforts in using genetic biomarkers to direct care of PanNET patients should take into account genetic heterogeneity of certain loci^31^: While *MEN1* aberrations are to be expected to be shared among all tumour cells, remaining PanNET driver events may indeed exhibit relevant heterogeneity that cannot be fully characterized using archived tissue or even a fresh tumour biopsy. We recommend that timely tissue acquisition should be required for includion in biomarker driven trials, and that longitudinal samples are studied to fully describe how new PanNET therapies may shape the biology of this disease. As for today, those PanNET patients that experience high grade progression after alkylating chemotherapy could be considered for TMB analysis, especially in countries where this result may provide additional treatment options. Finally our data strongly emphasize the need for novel and emerging methods, including blood based methods such as liquid biopsy as well as functional imaging that may both be able to provide real-time whole-body characterization of metastatic PanNET.

## Online Methods

### Ethics Statement

Ethical approval was granted through DNRs 2018/401, 2015/544/2 and 2012/422. This study was conducted accordingly to the Helsinki declaration.

### Discovery Cohort

Four different tissue collections at the Departments of Endocrine Oncology, Endocrine Surgery and Pathology as well as the Uppsala “U-CAN” collection^32^ were scrutinised to identify cases fulfilling the following criteria: patients with frozen panNET tissue samples available from metachronous time points. At least one tumour sample had to be from a metastatic lesion. Finally, each case had to have tissue or DNA from non-tumoural cells available. From the identified patients up to 3 tumour samples were acquired from each unique tumour lesion depending on availability. Tissue sections for histopathology were subjected to haematoxylin and eosin staining as well as MIB-1 immunostaining before examination accordingly to ENETS/WHO criteria by two expert endocrine pathologists (JB and CCJ)^33^.

### Pan-molecular characterisation of multi-region tumour samples

Tumour purity cut-off were set to 50% (specimens from surgical resections) and 30% (core needle biopsy) to proceed with nucleic acid extraction. The experimental strategy is overviewed in Figure 1. Samples were prepared so that DNA and RNA were obtained from the same tissue sections (surgical samples) or from neighbouring sections (core needle). All samples were subjected to pan-genomic characterisation with whole genome (surgical samples and corresponding non-tumoural tissue) or exome (core needle biopsies and corresponding non-tumoural tissue) sequencing. Further, surgical samples also underwent total RNA sequencing and where characterised using the Illumina EPIC DNA methylation array.

### Bioinformatics analysis

We aimed to perform a comprehensive characterisation of PanNET biology with particular focus on tumour intra- and inter heterogeneity. Experimental methods are described in full detail in the supplementary appendix. Briefly, mutation detection was performed using the Sarek pipeline, copy number analysis with ASCAT and Sclust, while RNA-seq analysis was performed using the nf-core-rnaseq pipeline and DNA methylation array data analysed using minfi. To put findings into perspective we compared all samples within the discovery cohort to each other. When appropriate we also analysed all samples within the discovery cohort from a pan-cancer perspective using data from the Cancer Genome Atlas (TCGA)^34^ as well as a separate cohort of gastroenteropancreatic neuroendocrine tumours^35^.

### Validation cohorts

Three validation experiments using different cohorts were conducted: In validation cohort 1 metastatic PanNET patients treated at Uppsala University Hospital with tumour samples available after treatment with alkylating chemotherapy were studied using targeted next generation sequencing with the Genomics Medicine Sweden (GMS)560 panel. In validation cohort 2 we aimed to describe the presence of hypermutation and lesions in *MSH2, MSH6, MLH1,* or *PMS2* in an unselected PanNET cohort by using a public DNA sequencing dataset of primary and metastatic PanNETs: American Association for Cancer Research Project Genomics Evidence Neoplasia Information Exchange (GENIE, https://genie.cbioportal.org)^23–25^. Finally, in validation cohort 3 we aimed to describe the mutational landscape of lynch syndrome associated PanNETs, such patients with tumour tissue available at Uppsala University Hospital were identified. Here tissues were characterized by Whole Genome Sequencing using the same approach as previously described.

### Statistical Analyses

Nominal data were presented as number of patients and percentages. Groups were compared by using Odds Ratio and Fisher’s exact test through Prism 10.1 (GraphPad Software Inc). P values <0.05 were considered as statistically significant. Statistical analyses of methylation and gene expression data were performed in R as described in the Supplementary Methods.

## Acknowledgements

The authors acknowledge the important contributions made by the included patients as well as all collaborates, funders and those maintaining research infrastructe:

Joakim Cronas research position is funded by Cancerfonden. This work was supported by grants from Cancerfonden, the European Neuroendocrine Tumor Society, Lions Cancerforsknignsfond Uppsala, Åke Wibergs Stiftelse and Söderbergs stiftelser.

Sequencing was performed by the SNP&SEQ Technology Platform in Uppsala. The facility is part of the National Genomics Infrastructure (NGI) Sweden and Science for Life Laboratory. The SNP&SEQ Platform is supported by the Swedish Research Council and the Knut and Alice Wallenberg Foundation.

Part of the material was collected with support of U-CAN, through Uppsala Biobank and the Department of Clinical Pathology, Uppsala University Hospital.

The computations and data handling were enabled by resources in project sens2019036 provided by the National Academic Infrastructure for Supercomputing in Sweden (NAISS) at UPPMAX, funded by the Swedish Research Council through grant agreement no. 2022-06725.

The authors acknowledge Clinical Genomics Uppsala, Science for Life Laboratory, Department of Immunology, Genetics and Pathology, Uppsala University, Sweden for providing assistance with Exome Sequencing and Targeted Sequencing (GMS560) including sample preparation, library preparation, bioinformatics and data interpretation.

This work was supported, by the Intramural Research Program of the National Institutes of Health, Eunice Kennedy Shriver National Institute of Child Health and Human Development. The authors would also like to acknowledge the American Association for Cancer Research and its financial and material support in the development of the AACR Project GENIE repository, as well as members of the consortium for their commitment to data sharing.

## Data availability

Access to the sensitive raw data can be requested through https://www.u-can.uu.se/. All additional data can be obtained from the corresponding author upon reasonable request.

## Code availability

The data analyses presented in this paper were performed using publicly available software as detailed in the Methods and Supplementary methods. Key scripts to reproduce figures and analyses have been deposited in a github repository (https://github.com/sabackman/PanNET_Evolution).

## Supplementary figure legends

Throughout supplementary figures PanNET samples were annoteted for location (P, Primary tumour; M, metastasis), Tumour lesion number (1-4) and Sample number from each individual tumour lesion (a-c).

Supplementary figure 1) Summary of treatment timelines and sample collection for patients included in the discovery cohort. Abbreviations: α-INF, Alfa-interferon; PRRT, Lu177-DOTATATE; SSA, Somatostatin; STZ: Streptozotocin analogue. α-INF: Alfa-interferon

Supplementary figure 2 A-F) Copy number profiles of samples from the discovery cohort.

Supplementary figure 3 A-F) Raw phylogenetic trees based on small mutation data for samples from the discovery cohort.

Supplementary figure 4) DNA methylation heatmaps and phyloepigenetic trees for patients PanNET01 (A-B), PanNET02 (C-D), PanNET0 (E-F) and PanNET04 (G-H). In the phyloepigenetic trees blue branches denote primary tumour samples and red branches metastatic lesions. Follow-up samples are marked in grey.

Supplementary figure 5) DNA methylation heatmap of all samples with available data. Samples cluster based on patient of origin.

Supplementary figure 6 a) Gene ontologies enriched among genes upregulated in follow-up samples from PanNET03. b) Gene ontologies enriched among genes downregulated in follow-up samples from PanNET03. c) Gene expression heatmap of all fresh-frozen samples. Supplementary figure 7) Clone distribution plots for patients PanNET01; a) PanNET02, b) and c) PanNET03. In PanNET01, the metastases share a clone (Clone 3) not present in primary tumours. In PanNET02, the primary tumours share Clone 2, not present in the metastatic sample potentially suggesting early metastatic seeding. In PanNET04 P1b/P1c share a clone with M2a/M2b, while P1a share a clone with M1a/M1b while M3 does not contain either of these clones, suggestive of multiple separate metastatic seeding events. Abbreviations: CCF, Cancer cell fraction.

Supplementary figure 8 a) Estimates used for deciding the number of mutation signatures to use for *de novo* mutation signature detection. Based on this plot extractions of 2-4 signatures were trialled. b-g) Mutation signatures and contributions using NMF extraction of 2 (B-C), 3 (D-E) and 4 (F-G) signatures. H) Estimated contributions of COSMIC v3 cancer mutation signatures identifies SBS11 as a strong contributor in the metastatic samples from PanNET03. I) Cosine similarity between raw and reconstructed mutation patterns related to (H).

Supplementary figure 9) Mutations shared between all metastatic samples from PanNET03 have a similar mutation signature to those found in the primary tumour samples, while mutations shared by M4a and M4b, as well as those shared by M1-M3 are dominated by the C>T SBS11-like signature.

Supplementary figure 10) DNA methylation difference between baseline and follow-up samples is greater in non-CpG-island sites.

Supplementary figure 11) Pan-cancer clustering of Immune function enrichment in a cohort of 24 tumour types. Abbreviations: GEPNET, Gastroenteropancreatic neuroendocrine tumours; PCPG, Pheochromocytoma and Paraganglioma; PanNET, Pancreatic neuroendocrine tumour and TCGA, the Cancer Genome Atlas.

Supplementary figure 12) Clustering of immune gene set enrichments (GSVA) across pancreatic tumour samples identified 3 clusters.

Supplementary figure 13) Genomic characteristics of 574 pancreatic NETs in the AACR GENIE cohort, a) 278 primary tumours, c) 289 metastases, and d) 8 *MSH6*-mutated metastases.

Supplementary figure 14) No evidence of significant contribution of SBS11 in a PanNET from a patient with Lynch syndrome.

Supplementary figure 15) Fraction of heterozygous SNPs with the same allele retained on chromosomes affected by cnLOH events in (A) PanNET03 and (B) PanNET04. For PanNET03 chromsomes 1-3, 6, 8-11, 15-16 and 21-22 were included. For PanNET04 chromosomes 1-3, 6, 8, 10-11, 15-16, 18 and 21-22 were included. Abbreviations: LOH, Loss of heterozygosity.

