## Supplementary materials&Methods for "The Evolutionary History of Metastatic Pancreatic Neuroendocrine Tumours Reveals a Therapy Driven Route to High-Grade Transformation"

**Supplementary methods**

Table 1. Pan-Cancer cohort

| Cohort | Cohort, full name | Ref |
| --- | --- | --- |
| **BLCA** | Bladder urothelial carcinoma | ^1,2^ |
| **BRCA** | Breast invasive carcinoma | ^3^ |
| **CESC** | Cervical squamous cell carcinoma and Endocervical adenocarcinoma | ^4^ |
| **COAD** | Colon adenocarcinoma | ^5^ |
| **GBM** | Glioblastoma multiforme | ^6,7^ |
| **HNSC** | Head and neck squamous cell carcinoma | ^8^ |
| **KICH** | Kidney chromophobe | ^9^ |
| **KIRC** | Kidney renal clear cell carcinoma | ^10^ |
| **KIRP** | Kidney renal papillary cell carcinoma | ^11^ |
| **LGG** | Brain Lower Grade Glioma | ^6,12^ |
| **LIHC** | Liver hepatocellular carcinoma | ^13^ |
| **LUAD** | Lung adenocarcinoma | ^14^ |
| **OV** | Ovarian serous cystadenocarcinoma | ^15^ |
| **PAAD** | Pancreatic adenocarcinoma | ^16^ |
| **PCPG** | Pheochromocytoma and paraganglioma | ^17^ |
| **PRAD** | Prostate adenocarcinoma | ^18^ |
| **READ** | Rectum adenocarcinoma | ^5^ |
| **SARC** | Sarcoma | ^19^ |
| **SKCM** | Skin cutaneous melanoma | ^20^ |
| **STAD** | Stomach adenocarcinoma | ^21^ |
| **THCA** | Thyroid carcinoma | ^22^ |
| **UCEC** | Uterine corpus endometrial Carcinoma | ^23^ |
| **UVM** | Uveal melanoma | ^24^ |
| **GET-NETs** | Gastroenteropancreatic Neuroendocrine Tumours | ^25^ |

Validation cohorts

Validation cohort 1: we screened all PanNET patients treated at the Department of Endocrine Oncology, Akademiska Sjukhuset, Uppsala, Sweden since 1980. Patients with longitudinal samples were selected. We identified those that had either surgical resection or core-needle biopsy samples taken after treatment with alkylating agents Streptozotocin and/or Temozolomide. For those patients we aimed to identify one tumour sample per patient. In the case of multiple samples available, the most recently acquired sample after discontinuation of temozolomide/streptozotocin was prioritized. Ki67 values were obtained for each case from the electronic medical records that had employed standard WHO/ENETS criteria^26^.

Validation cohort 2: The American Association for Cancer Research Project Genomics Evidence Neoplasia Information Exchange Cohort v13.0-public was accessed using cBioportal (<https://genie.cbioportal.org>) on 2023-09-14^27-29^. We selected patients with PanNET having with information on mutation count and mismatch repair mutation status and stratified by sample type: primary tumour and metastatic lesion.

Validation cohort 3: we screened all PanNET patients managed at the Departments of Endocrine oncology and Endocrine Surgery since 1980. Patients with clinically confirmed Lynch syndrome were identified, and we scrutinized for available tumour tissue and matched non-tumoural tissue. The obtained specimens underwent whole genome sequencing following the same workflow as in the discovery cohort.

Tissue preparation

Tumour tissues were collected from surgical specimens or from core needle biopsies that had been snap frozen in liquid nitrogen within 2 hours after retrieval and stored in in -80°C. The tissue was cut using a cryostat: For samples acquired from surgical resection, approximately ten 20µm thick sections were collected for DNA and RNA extraction. Two 6 µm sections was taken before and after the tissue collected for nucleic acid extraction to be used for histopathology analyses. For core needle biopsies fifteen 4 µm sections were taken for DNA extraction with another fifteen 4 µm sections were taken for RNA extraction. In addition, two 4 µm sections, one at the top and one at the bottom were acquired for histopathology. Tissue sections for histopathology were subjected to haematoxylin and eosin staining as well as MIB-1 immunostaining before examination by two expert endocrine pathologists (JB and CCJ). First, PanNET diagnosis was confirmed with estimation of PanNET cell fraction. For the included samples analysis of Ki-67 index was performed: Section 6 µm were immersed in 10% hydrogen peroxide solution for 15 minutes, then subsequently treated with avidin and biotin (Vector Laboratories, Burlingame, USA) for 30 minutes. The sections were then preincubated with 10% normal horse serum for 15 minutes, after which the primary antibody (concentration 1:100) was applied for 2 hours. For detection, biotinylated HRP-conjugated horse anti-mouse secondary antibodies were used, 1:200 (Vector Laboratories, Burlingame, USA). The sections were developed with DAB (Vector Laboratories, Burlingame, USA) and counterstained with hematoxylin. The haematoxylin & eosin as well as MIB-1 immunostained slides were analysed to determine Ki-67 index and PanNET grade. Ki67 was analysed accordingly to WHO/ENETS criteria by analysing 2000 cells in hot spots^26^.

Nucleic acids were extracted accordingly to the following protocol. Surgical samples: Genomic DNA and RNA was isolated using AllPrep DNA/RNA/miRNA Universal Kit (Qiagen, Hilden, Germany) according to the manufacturer’s instructions.

Core needle biopsies: Genomic DNA was isolated using QIAamp Fast DNA Tissue Kit (Qiagen, Hilden, Germany) and the DNA quantity and quality (DIN value) were assessed using Genomic DNA Screen Tapes on a 2200 TapeStation system (Agilent, Santa Clara, CA, USA). Genomic DNA were also extracted from whole blood using QIAamp DNA Blood Mini Kit (Qiagen, Hilden, Germany). The DNA quantity and quality (DIN value) were assessed using Genomic DNA Screen Tapes on a 2200 TapeStation system (Agilent, Santa Clara, CA, USA).

Whole genome sequencing

All samples from surgical resection and the corresponding non-tumoural tissue were subjected to whole genome sequencing at Science for Life Laboratory, Uppsala Node ([https://snpseq.medsci.uu.se](https://snpseq.medsci.uu.se/)). Sequencing libraries were prepared from 1μg DNA using the TruSeq PCRfree DNA sample preparation kit (cat# 20015962, Illumina Inc.) and unique dual indexes (cat#20022370, Illumina Inc.), targeting an insert size of 350bp. The library preparation was performed according to the manufacturers’ instructions (guide#1000000039279). Paired-end 150bp read length, NovaSeq 6000 system, SP flowcell and v1 sequencing chemistry.

Whole exome sequencing

All samples from core needle biopsies and the corresponding non-tumoural tissue were subjected to exome sequencing at Clinical Genomics and SNP&SEQ Technology Platform, Science for Life Laboratory, Uppsala, Sweden. Exome libraries were prepared using the Twist Library Preparation Kit (Mechanical Fragmentation) and target enrichment protocol (Twist Bioscience, San Francisco, CA, USA), using the Twist core exome panel. The libraries were paired-end sequenced (100 cycles) on two S1 flow cells using the NovaSeq 6000 system and v1 sequencing chemistry (Illumina San Diego, CA, USA).

DNA methylation array

Methylation profiling by genotyping was performed with the Illuminas Infinium assay (MethylationEPIC_v-1-0) by Science for Life Laboratory, Uppsala Node ([https://snpseq.medsci.uu.se](https://snpseq.medsci.uu.se/)). Bisulfite conversion was performed using the EZ DNA MethylationTM Kit from Zymo Research. Product No: D5004 with 250 ng of DNA per sample. The bisulfite converted DNA was eluted in 15μl according to the manufacturer’s protocol, evaporated to a volume of <4μl, and used for methylation analysis using the Illumina Methylation EPIC array. The resulting data was processed using the minfi R-package (1.44.0) ^30^, using preprocessIllumina as the normalization function. Heatmaps were generated based on the 8640 (1%) most variable probes as measured by standard deviation. The statistical analysis presented in Supplementary Figure 10 was performed using the t-test as implemented in R version 4.2.2.

**Bioinformatics analyses**

Scripts for reproducing key results and figures are made available in a dedicated github repository: <https://github.com/sabackman/PanNET_Evolution>

Bioinformatics analysis of DNA sequencing

Generated BAM files were marked for duplicate reads. Subsequently the nf-core/Sarek^31^ pipeline (version 2.6.1 with minor modifications) was used to perform Base Quality Score

Coverage data for exome sequencing samples was calculated using mosdepth^32^. Recalibration, variant calling with MSISensor^33^, Mutect2^34^ and Manta, as well as variant annotation using VEP and snpEff. For variant calling with Mutect2, a somatic Panel of Normals generated from 19 germline samples from patients with endocrine tumours sequenced in the same run was used. Only variants passing all Mutect2 filters were included in the final analysis. For the exome sequencing samples, a generic Panel of Normals provided in the GATK resource bundle was used. Germline variants were called using GATK HaplotypeCaller. Throughout the analysis, non-synonymous variants were defined as splice site/splice region/start lost/stop lost/stop gained/missense/frameshift variant and inframe insertions and deletions. Tumour mutation burden was calculated as the number of called somatic variants passing filters, divided by cumulative the length of the genomic regions in which mutations were called (i.e. the target regions input to Mutect2) multiplied by 10^6^. Based on empirical data tumour mutation burden results was categorized as ≥50 (hypermutated) or <50 (non-hypermutated). Variants in known cancer associated genes/DNA reparation genes with a population frequency less than 2% were extracted and manually reviewed.

Excluding cancer-associated pathogenic germline mutations

HaplotypeCaller variant calls affecting the protein coding sequence of a set of 96 known cancer-associated genes were manually reviewed, taking into account population frequency (gnomAD) and previous reports on pathogenicity (ClinVar).

**Table 2. Genes screened for germline pathogenic variants**

| *PALB2* | *BRCA2* | *PMS1* | *MITF* | *DICER1* |
| --- | --- | --- | --- | --- |
| *MUTYH* | *ATM* | *FANCF* | *KIT* | *SDHC* |
| *MEN1* | *APC* | *ERCC2* | *FH* | *MSH2* |
| *CDKN1B* | *PTCH1* | *DIS3L2* | *FANCG* | *HOXB13* |
| *CHEK2* | *MPL* | *XPA* | *FANCE* | *ERCC3* |
| *TSC2* | *MET* | *WAS* | *FANCD2* | *RAD51D* |
| *WRN* | *FANCM* | *TSHR* | *FANCC* | *PRKAR1A* |
| *FANCL* | *ERCC4* | *TSC1* | *EXT2* | *CDKN2A* |
| *ALK* | *BARD1* | *TP53* | *EGFR* | *CDK4* |
| *RTEL1* | *BAP1* | *SUFU* | *CTNNA1* | *CDH1* |
| *BLM* | *PRF1* | *SDHD* | *BRIP1* | *BMPR1A* |
| *VHL* | *PMS2* | *SDHB* | *XPC* | *PHOX2B* |
| *TERT* | *GATA2* | *SDHA* | *STK11* | *MAX* |
| *RET* | *FAM175A* | *RECQL4* | *SMAD4* | *XRCC2* |
| *RAD54L* | *ERCC5* | *RAD50* | *RB1* | *SBDS* |
| *POLE* | *BUB1B* | *NTRK1* | *NF1* | *FANCB* |
| *POLD1* | *BRCA1* | *NF2* | *MSH6* |  |
| *KLLN* | *ATR* | *NEK1* | *MLH1* |  |
| *FANCI* | *SLX4* | *NBN* | *EXT1* |  |
| *FANCA* | *RAD51C* | *MRE11A* | *EPCAM* |  |

Mutation signature analysis

Mutation signature analysis was performed using the MutationalPatterns R package^35^. De novo mutation signature deconstruction was performed on the above described variant callset using Non-negative matrix factorisation as implemented in the MutationalPatterns nmf function using the brunet method and 100 runs. A manual review of the results was performed prior to deciding the number of signatures to extract. The variants were also fitted to established COSMIC mutation signatures using the fit_to_signatures function with the standard parameters.

Copy number analyses

Whole genome sequencing samples were subjected to copy number analyses using Sclust^36^ and ASCAT^37,38^. Cases were the two tools reported discordant ploidy/purity combinations were manually reviewed and in most cases the Sclust analyses was rerun with forced policy. The samples from one patient (PanNET01) had very variable ploidy estimates in both the ASCAT and Sclust analyses. For this sample the subsequent evolutionary analyses were run using several different ploidy combinations. Exome sequencing samples were subjected to copy number analyses using Sclust, using the appropriate settings for exome sequencing.

Evolutionary analyses

A binary matrix was generated for each patient, indicating the presence of absence of each somatic mutation (SNV/small indel) in each sample. The R *dist* function was used to generate a distance matrix. The *fastme.bal* function from the *ape*^39^ R package was used to generate phylogenetic trees.

For DNA methylation data the 1% most variable probes as measured by range were extracted, and a distance matrix constructed using the *dist* function, followed by construction of phylogenetic trees using the fastme.bal function from the *ape* package. For comparison of tree topologies we used the RF.dist function from the *phangorn*^40^ R package.

Subclonal reconstruction was performed using PyClone-VI (0.1.0)^41^. Mutations on autosomal chromosomes were included if the locus had a read count of at least 40 in each tumour sample, and at least 20 in the paired normal. Moreover, the mutation had to pass all MuTect2 filters and have an MMQ score of at least 50 in all samples where it was called. Read counts for mutations not called in a given sample were retrieved using samtools mpileup and a custom wrapper script. Mutations located in problematic genome regions (hg38-blacklist.v2.bed)^42^, or in low-complexity regions (retrieved from Gnomad) were excluded. Mutations were also excluded if they were present with an allele frequency of more than 0.0001 in Gnomad. Copy number calls were generated using Sclust (1.1)^36^. Mutations were excluded if copy number calls for the region were missing in any sample, if the mutation overlapped a subclonal copy number aberration, or if either allele was present in more than two copies. Mutation copy number was modelled as $Mutated CN=AF*\frac{Purity*Total CN+\left( 1-purity \right)*2}{purity}$. Heuristically mutations with a modelled copy number > 1.5 were assigned a copy number of 2, while the rest were assigned a copy number of 1. Simulations suggested that the alternative approach of assigning mutation copy number using maximum likelihood estimation would have a minor effect on the results, altering the assigned copy number for on average 0.017% of the variants (range 0-0.042%). Mutations were clustered using PyClone-VI with a beta-binomial model and 100 random restarts.

In the case of PanNET03, where the metastatic samples exhibited massive hypermutation, clonal reconstruction using the full dataset was computationally intractable. For this sample mutations present in at least one of the primary tumours were included, and 5% of mutations present only in metastatic samples were included at random. Similar to Burdett et al^43^. We excluded subclonal clusters that had a cancer cell fraction (CCF) within a 10% range in all samples, and we also excluded clusters with less than 20 mutations.

Timing of *ATRX/DAXX* mutations in relation to copy number alterations

The mutation copy number for the *ATRX* and *DAXX* mutations was modelled using a binomial distribution, and the binomial test was used to test whether the mutation occurred prior to or after loss of heterozygosity (LOH) and genome doubling, i.e. whether it was present on one or two chromosomes.

In order to determine whether LOH events retained the same physical chromosome across samples, read counts for heterozygous loci were extracted using samtools mpileup. Due to computational constrains, 10% of such sites were included at random. The allele with the highest read count on chromosomes with LOH was considered to retained. For chromosomes affected by LOH, we computed the fraction of SNPs with the same retained allele between all pairs of samples, expecting this to be close to 1 if the same physical chromosome is retained, and close to 0 if different physical chromosomes are retained in different samples.

Total RNA sequencing

RNA sequencing was performed at Science for Life Laboratory, Uppsala Node ([https://snpseq.medsci.uu.se](https://snpseq.medsci.uu.se/)). Sequencing libraries were prepared from 1μg total RNA using the TruSeq stranded total RNA library preparation kit with RiboZero Gold treatment (cat# 20020598/9, Illumina Inc.). The library preparation was performed according to the manufacturers’ protocol (# 1000000040499). Sequencing was performed using NovaSeq S4 flowcell, paired-end 150bp read length, v1 sequencing chemistry.

One sample had a deviating expression profile thought to be caused by RNA degradation and was excluded from further analyses.

RNA-Seq data processing

The RNA-Seq data was analysed using the nf-core/rnaseq pipeline v 3.9^31^, in which reads were aligned using STAR^44^ and transcripts quantified using Salmon^45^. Differential expression analyses were performed using DeSeq2 (1.38.3)^46^. For the heatmap generation, the z-transformed normalized counts from the 5% of genes with the highest variation as measured by variance/mean was used. One sample, PanNET04 M2b, was excluded from this analysis due to signs of low library complexity. Gene ontology enrichment analyses were performed using the *enrichGO* function from the *clusterProfiler* (4.6.2)^47^ R package, including genes with an absolute log2 fold change greater than 1.5 and adjuster p-value < 0.05.

Immunotranscriptomic analyses

For the immunotranscriptomic analysis, the FASTQ-files were analysed using nf-core/rnaseq pipeline v 1.0 and raw counts from featureCounts were further analysed using CIBERSORT^48^ as previously described^49^. Briefly, raw RNA-Seq v2 data, containing a total of 8,631 tumor samples and processed through the HTseq-count pipeline, was obtained from the Genomic Data Commons portal. In addition, the transcriptomic profiles of three Gastroenteropancreatic neuroendocrine tumor subtypes were derived from a previously published study^25^. To ensure uniform analysis, raw counts data from all sources were pre-processed using the edgeR R package, resulting in log counts per million data for subsequent analysis^25^. Then we curated gene signatures from four established resources. These resources included the LM22 immune infiltration signature used in CIBERSORT, the LM7 immune infiltration signature developed by Tosolini and colleagues^50^, the ImSig signature for solid tumour immune infiltration by Nirmal and colleagues^51^, and the NanoString immune signature panel (accessible at https://www.nanostring.com). In total, 61 distinct gene sets were compiled from these sources. This geneset was used for single-sample gene set enrichment analysis (ssGSEA) implemented on the gene expression data of our combined tumour cohort. R package GSVA (Gene Set Variation Analysis) was used for this analysis. Furthermore, we conducted unsupervised clustering of ssGSEA scores across tumour samples, utilizing Spearman’s correlation coefficient as a similarity metric.

Genomics Medicine Sweden GMS560 gene panel sequencing

Targeted sequencing using the GMS560 gene panel was performed at Clinical Genomics Uppsala, Science for Life Laboratory. The GMS560 gene panel consists of a genomic DNA target capture panel module for comprehensive genomic profiling of solid tumours (comprehensively outlined at^52^). The design, optimization and validation are the result of a nationwide collaborative effort within the Genomic Medicine Sweden (GMS) Solid Tumour work package (https://genomicmedicine.se). Kapa Biosystems reagents (Wilmington, MA, USA) in combination with Twist Bioscience probes (Twist Bioscience, San Francisco, CA, USA) and reagents were used for library preparation and enrichment, respectively, following the manufacturers’ protocols with minor modifications. The libraries were paired-end sequenced (150 cycles) on a NextSeq 550 system (Illumina San Diego, CA, USA).

The signal profile of the GMS560 data from Validation cohort 1 was created using the SigProfiler tools. First, SigProfilerMatrixGenerator v1.2.19 was used to generate a single base substitution matrix of size 96 (SBS96) commonly used when describing cancer profiles^53^. Thereafter, the matrix was used by SigProfilerExtractor v1.1.22 to obtain an optimal set of signal profiles^54^. To find the optimal size of the profile set, the size of the number of profiles were varied from 1 to 10 using 100 replicates each, and then chosen based on the recommended value in the selection plot. The contribution of the profiles for each sample was finally calculated using the number of mutations associated with respective profile.

**Results**

What comes first; copynumber neutral (cnn)LOH or *ATRX/DAXX* mutation?

**Table 3. PanNET03 *DAXX***

| **Sample** | | **CN mutation** | **95% lower bound** | **95% upper bound** | **p-value After LOH** | **p-value before LOH** |
| --- | --- | --- | --- | --- | --- | --- |
| M1 | 1.692307692 | | 1.153489311 | 2.259445681 | 0.005712334 | 0.261836381 |
| M2 | 2.031063321 | | 1.572314171 | 2.442235496 | 1.93E-06 | 1 |
| M3a | 1.986928105 | | 1.659878586 | 2.200213619 | 8.72E-09 | 0.83602413 |
| M3b | 2.017510468 | | 1.688675033 | 2.295743451 | 3.73E-10 | 1 |
| M4a | 2.150537634 | | 1.461699513 | 2.930942652 | 0.000214789 | 0.702527985 |
| M4b | 1.714285714 | | 1.311635897 | 2.08506037 | 0.000181156 | 0.107546323 |

The estimated copy number of the mutated allele is close to 2, the binomial test suggests that the mutation occurred prior to cnnLOH in all samples.

**Table 4. PanNET03 *ATRX***

| **Sample** | **CN mutation** | **95% lower bound** | **95% upper bound** | **p-value After LOH** | **p-value before LOH** |
| --- | --- | --- | --- | --- | --- |
| P1a | 2.012870013 | 1.710551167 | 2.153142742 | 3.70E-09 | 1 |
| P1b | 2.117424242 | 1.802233257 | 2.298245575 | 1.84E-10 | 0.538643725 |
| P1c | 1.828282828 | 1.503558396 | 2.051514689 | 8.02E-07 | 0.131823077 |

Under the assumption that the X chromosome is present in two copies, the data suggest that the mutation occurred prior to LOH.

**Table 5. PanNET04 *DAXX***

| **Sample** | **CN mutation** | **95% lower bound** | **95% upper bound** | **p-value After LOH** | **p-value before LOH** |
| --- | --- | --- | --- | --- | --- |
| P1b | 2.040816327 | 1.60543401 | 2.404354566 | 5.01E-07 | 1 |
| M1a | 1.842105263 | 1.494518069 | 2.135654091 | 6.99E-07 | 0.290011977 |
| M1b | 1.872146119 | 1.50805565 | 2.184724638 | 6.44E-07 | 0.466826147 |
| M2a | 1.990049751 | 1.706390756 | 2.203556894 | 3.50E-11 | 0.87707207 |
| M2b | 1.902748414 | 1.564798023 | 2.135072463 | 2.63E-07 | 0.387329027 |
| P1c | 2.169981917 | 1.809129514 | 2.39421808 | 1.54E-09 | 0.346717461 |
| M1c | 2.024691358 | 1.662919344 | 2.307030877 | 1.55E-08 | 1 |
| P1a | 1.886792453 | 1.50423239 | 2.207021578 | 2.20E-06 | 0.530793326 |
| M3 | 1.860974275 | 1.411812163 | 2.29684711 | 3.73E-05 | 0.525685292 |

Validation cohort 3

Here 525 patients were screened and one PanNET patient having clinically confirmed Lynch syndrome was identified. A sample from the primary tumour as well as non-tumoural tissue was subjected to whole genome sequencing using the exact same methodology as that employed in the discovery cohort. 23 protein coding mutations corresponding to a tumour mutational burden of 1mut/Mb (not elevated) was discovered. No trace of mutation signature 11 could be discovered. MSIsensor was 0%
