## Supplementary figures for "The Evolutionary History of Metastatic Pancreatic Neuroendocrine Tumours Reveals a Therapy Driven Route to High-Grade Transformation"

**Supplementary Figure 01**

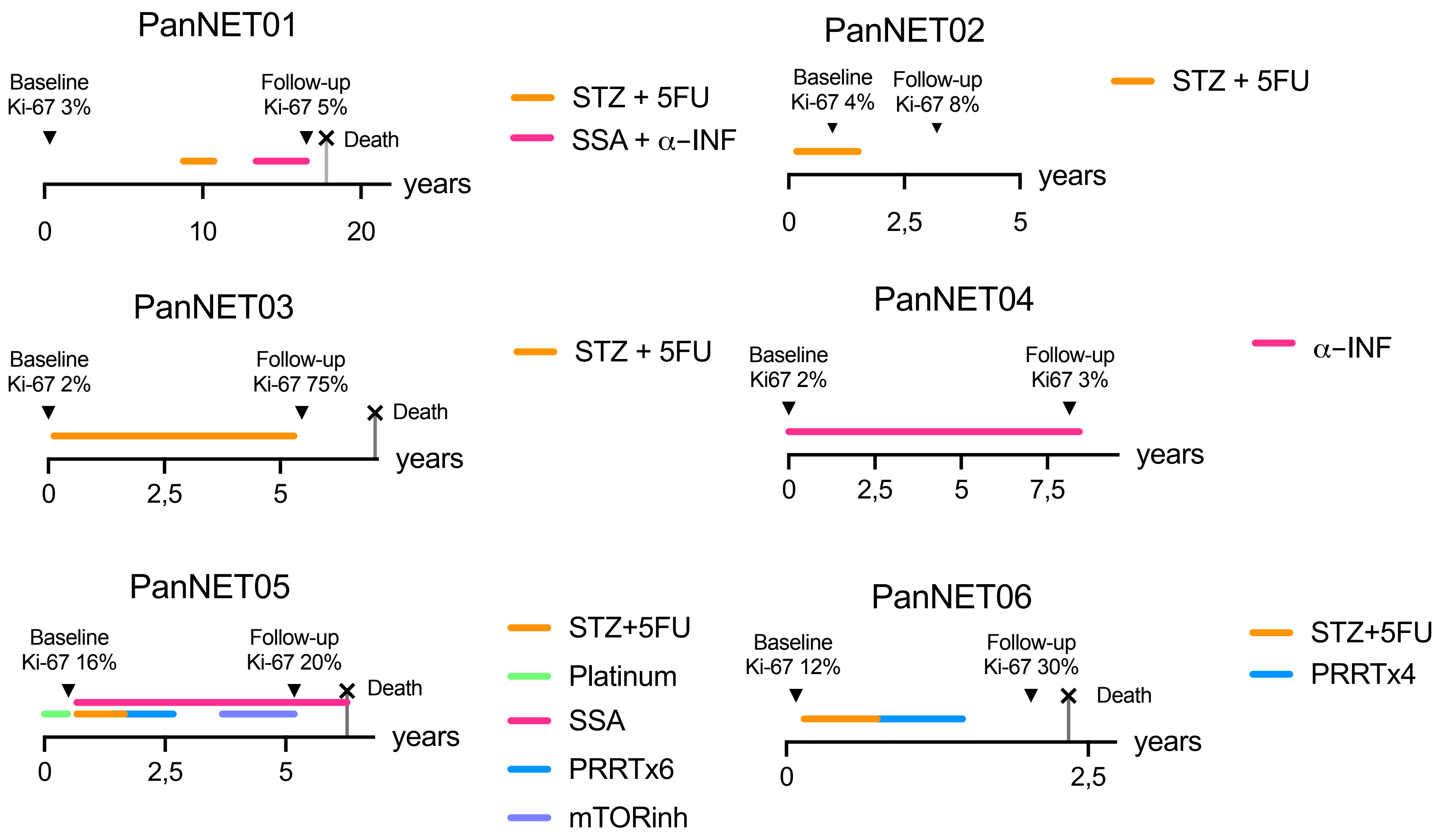

### Supplementary Figures 02 A-F

#### Supplementary Figure 02 A – PanNET01

##### Primary tumors

**P1a**

Ploidy: 2.01, aberrant cell fraction: 76%, goodness of fit: 97.5%

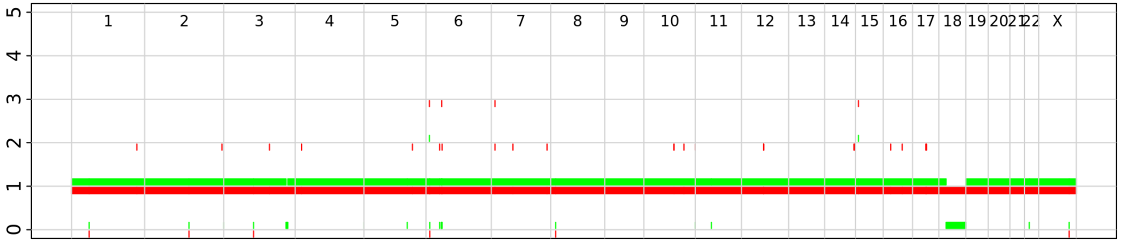


**P1b**

Ploidy: 4.01, aberrant cell fraction: 62%, goodness of fit: 91.6%

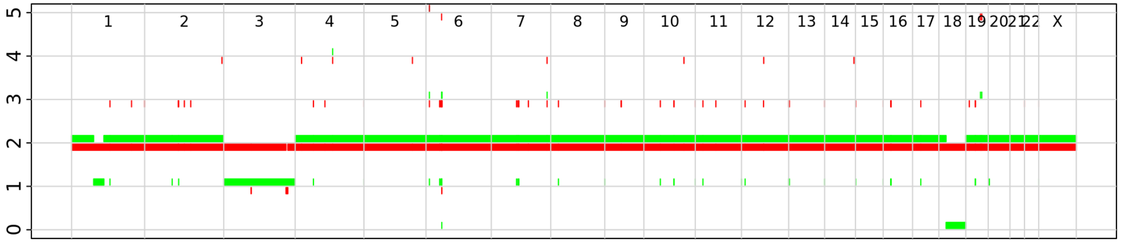


**P1c**

Ploidy: 3.98, aberrant cell fraction: 78%, goodness of fit: 92.3%

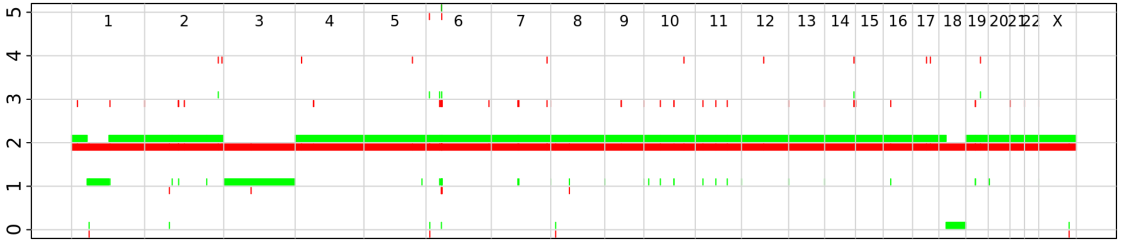


##### Metastases

**M1a**

Ploidy: 3.95, aberrant cell fraction: 78%, goodness of fit: 97.5%

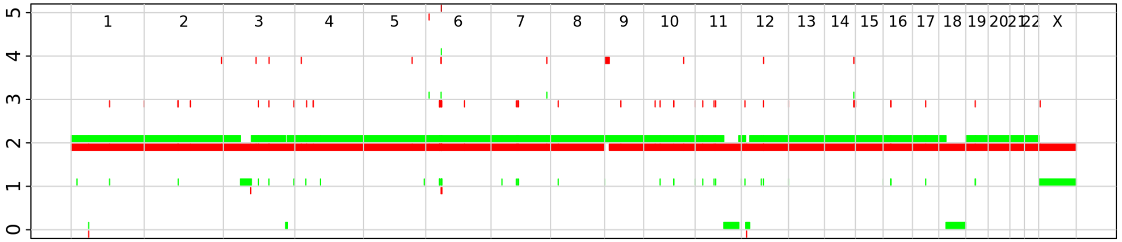


**M1b**

Ploidy: 1.92, aberrant cell fraction: 88%, goodness of fit: 97.1%

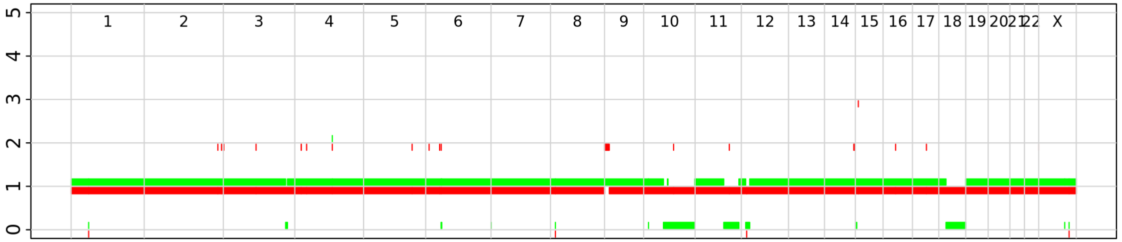


**M2**

Ploidy: 3.98, aberrant cell fraction: 80%, goodness of fit: 94.3%

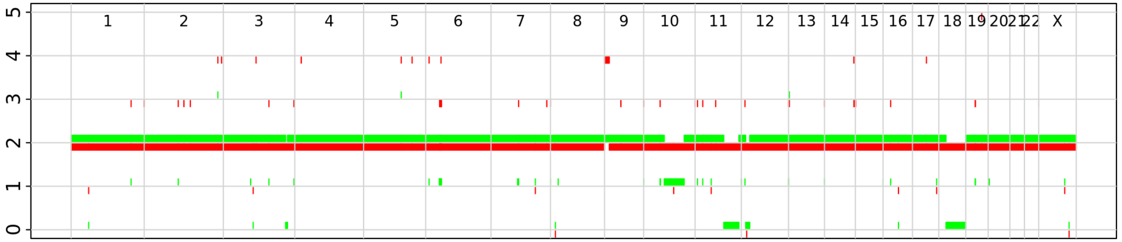

### Supplementary Figures 02 A-F

#### Supplementary Figure 02 B – PanNET02

##### Primary tumors

**P1a**

Ploidy: 1.94, aberrant cell fraction: 91%, goodness of fit: 99.2%

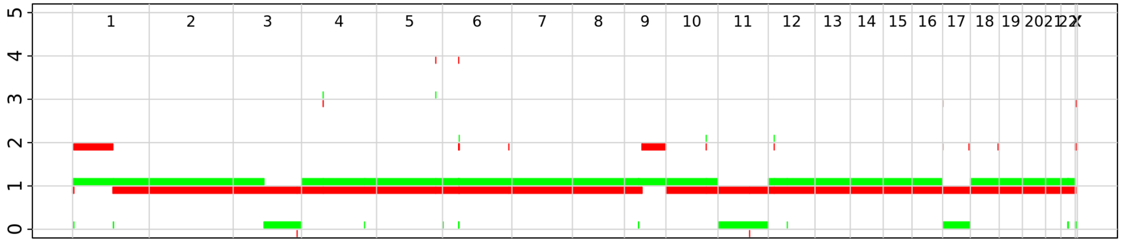


**P1b**

Ploidy: 1.89, aberrant cell fraction: 92%, goodness of fit: 98.9%

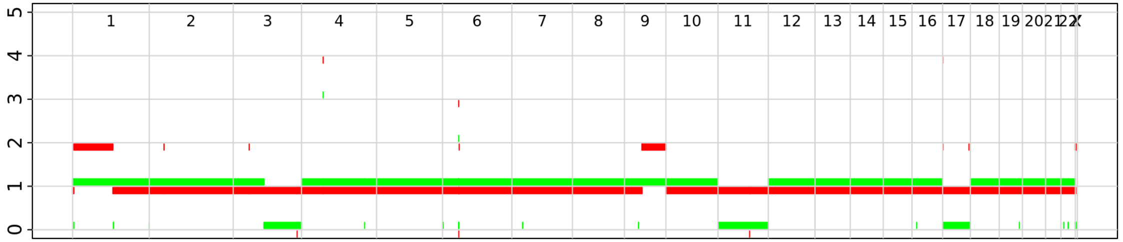


##### Metastases

**M1**

Ploidy: 1.98, aberrant cell fraction: 67%, goodness of fit: 98.2%

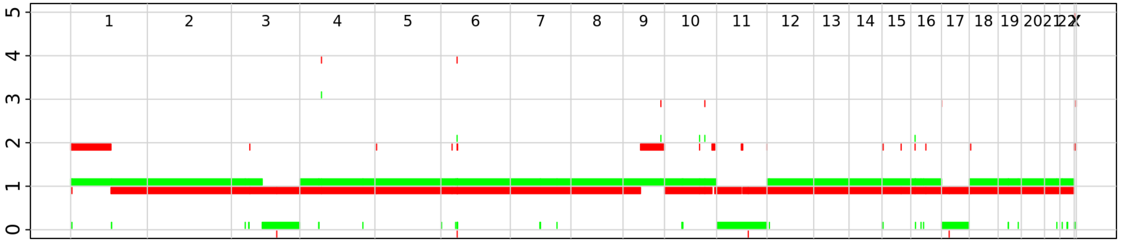

### Supplementary Figures 02 A-F

#### Supplementary Figures 02 C – PanNET03

#### Primary tumors

**P1a** Ploidy: 2.77, aberrant cell fraction: 84%, goodness of fit: 99.7%

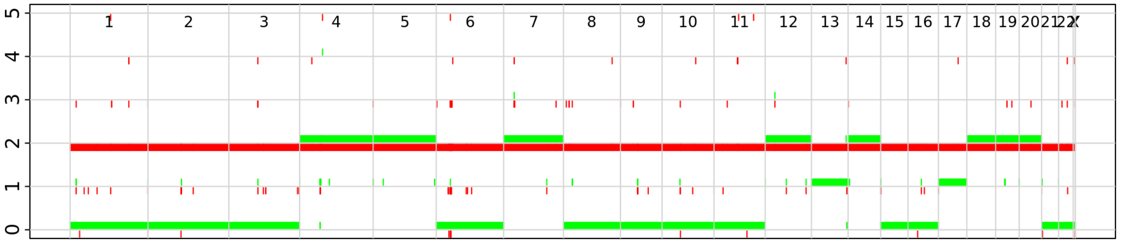


**P1b** Ploidy: 2.80, aberrant cell fraction: 72%, goodness of fit: 99.7%

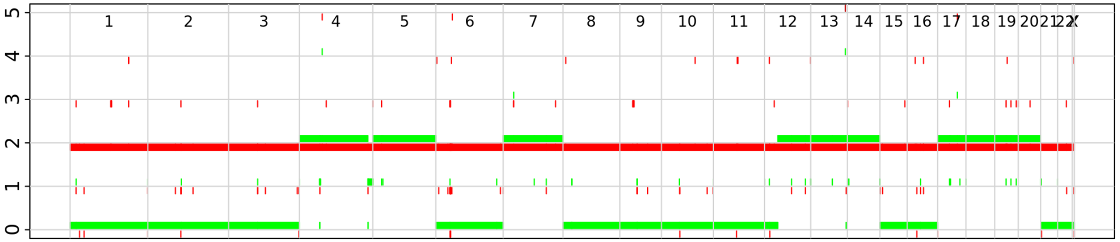


**P1c** Ploidy: 2.81, aberrant cell fraction: 81%, goodness of fit: 99.6%

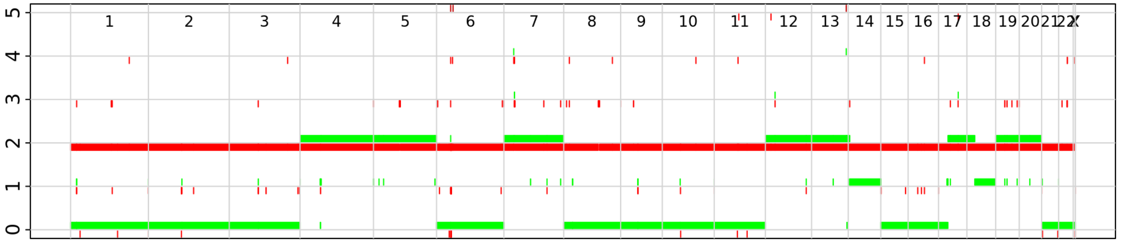


#### Metastases

**M1** Ploidy: 2.67, aberrant cell fraction: 52%, goodness of fit: 99.7%

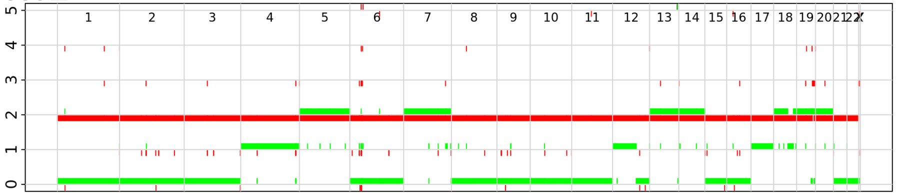


**M2** Ploidy: 2.58, aberrant cell fraction: 62%, goodness of fit: 99.3%

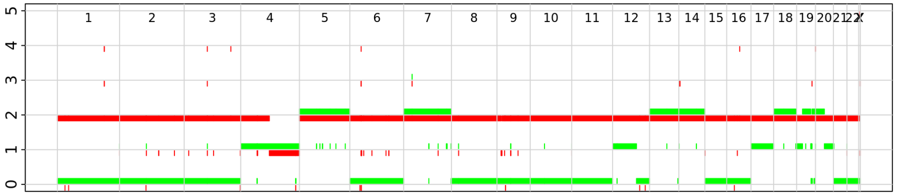


**M3a** Ploidy: 2.54, aberrant cell fraction: 85%, goodness of fit: 99.7%

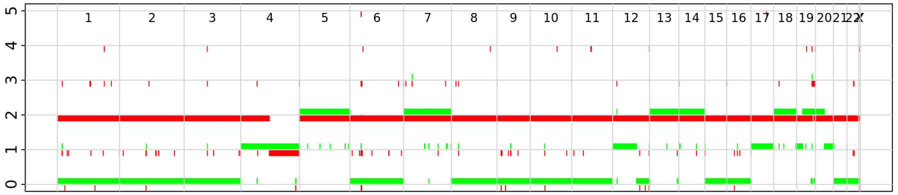


**M3b** Ploidy: 2.55, aberrant cell fraction: 71%, goodness of fit: 99.8%

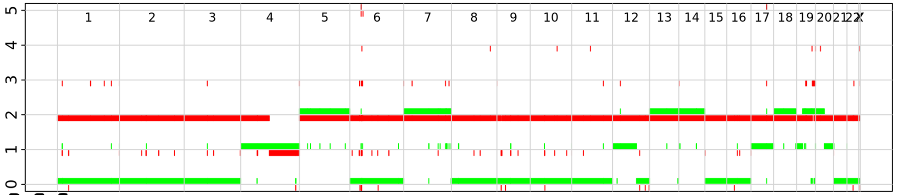


**M4a** Ploidy: 2.99, aberrant cell fraction: 31%, goodness of fit: 99.1%

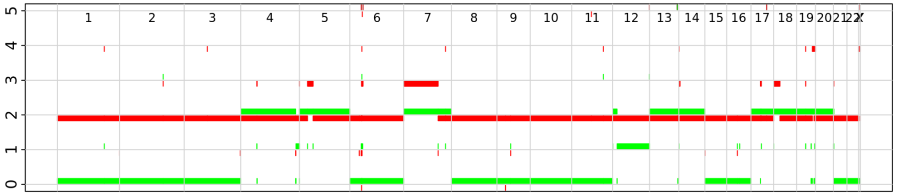


**M4b** Ploidy: 2.60, aberrant cell fraction: 70%, goodness of fit: 98.8%

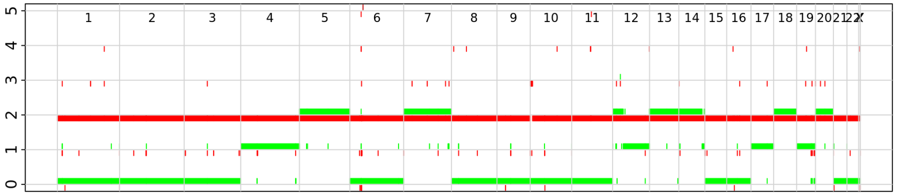

### Supplementary Figures 02 A-F

#### Supplementary Figure 02 D – PanNET04

##### Primary tumors

**P1a**

Ploidy: 2.79, aberrant cell fraction: 74%, goodness of fit: 99.6%

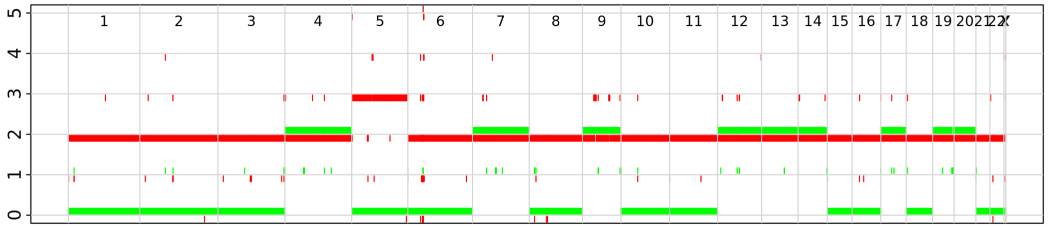

**P1b**

Ploidy: 2.76, aberrant cell fraction: 68%, goodness of fit: 99.7%

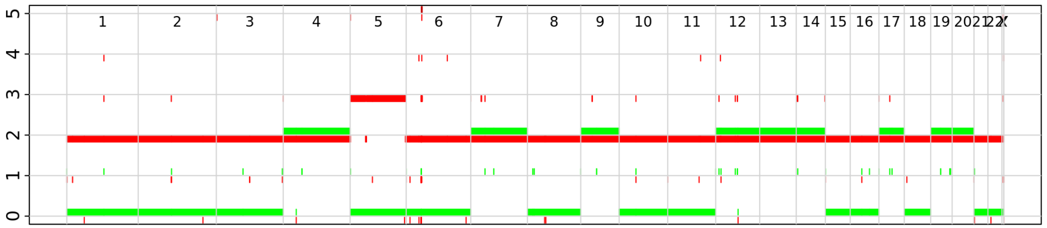

**P1c**

Ploidy: 2.80, aberrant cell fraction: 79%, goodness of fit: 99.8%

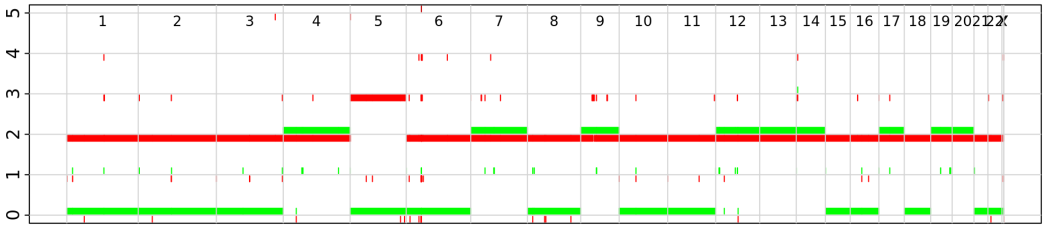

##### Metastases

**M1a**

Ploidy: 2.73, aberrant cell fraction: 76%, goodness of fit: 99.7%

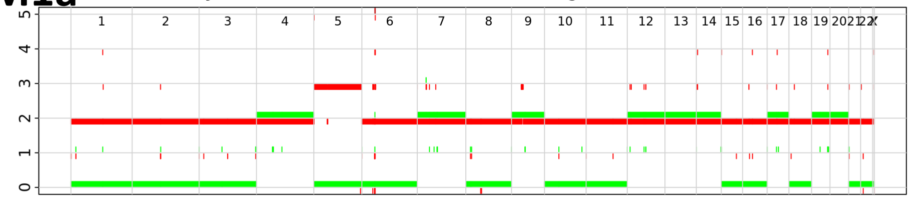

**M1b**

Ploidy: 2.78, aberrant cell fraction: 73%, goodness of fit: 99.7%

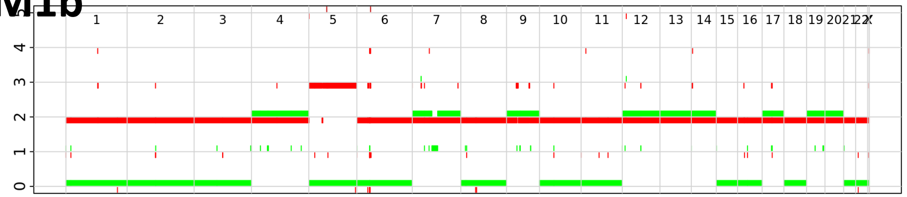

**M1c**

Ploidy: 2.78, aberrant cell fraction: 75%, goodness of fit: 99.7%

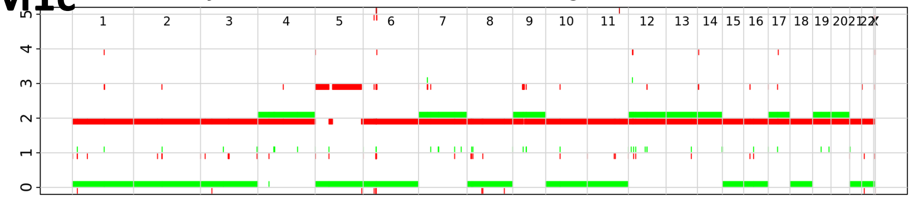

**M2a**

Ploidy: 2.75, aberrant cell fraction: 81%, goodness of fit: 99.7%

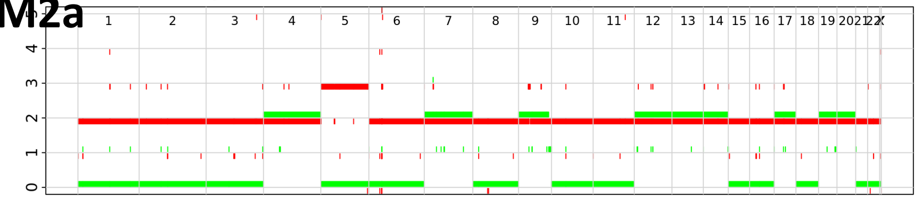

**M2b**

Ploidy: 2.82, aberrant cell fraction: 86%, goodness of fit: 99.7%

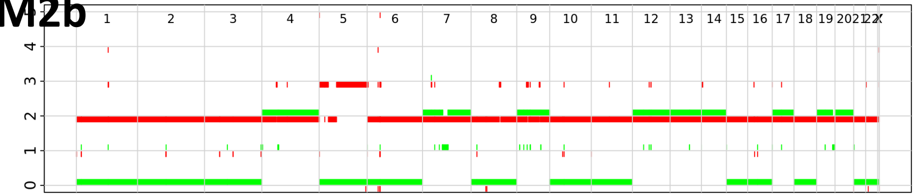

**M3**

Ploidy: 2.85, aberrant cell fraction: 58%, goodness of fit: 99.5%

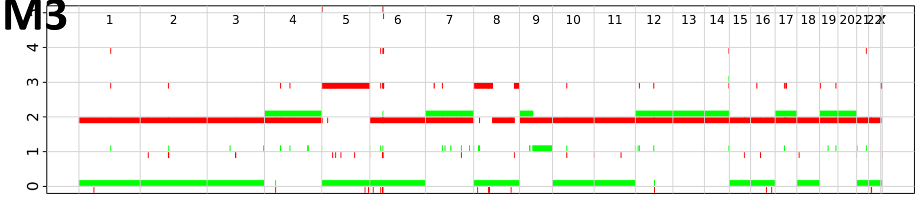

### Supplementary Figures 02 A-F

#### Supplementary Figure 02 E – PanNET05

M1a

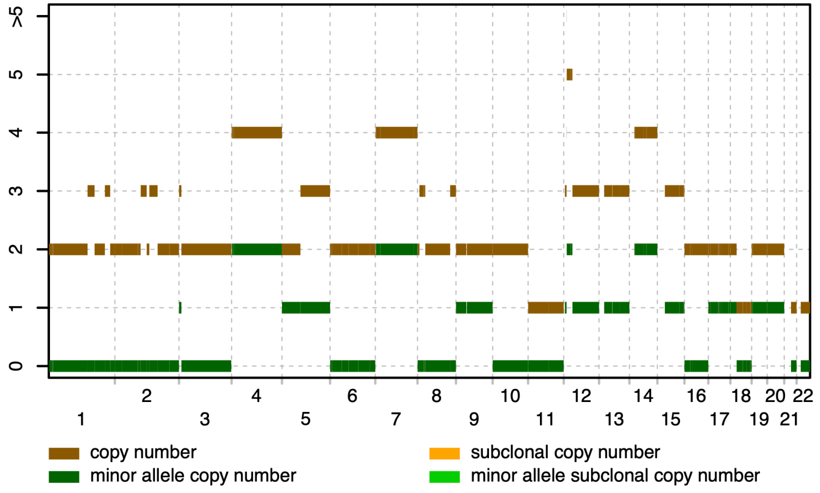


M2

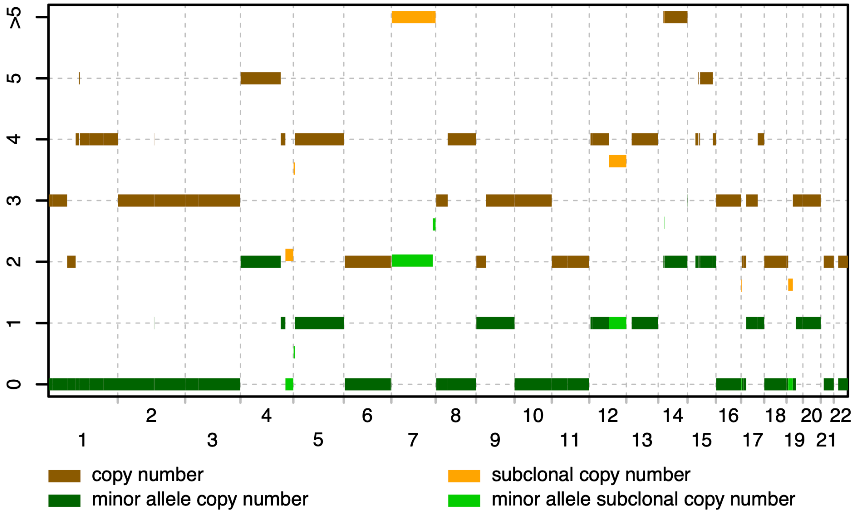


M1b

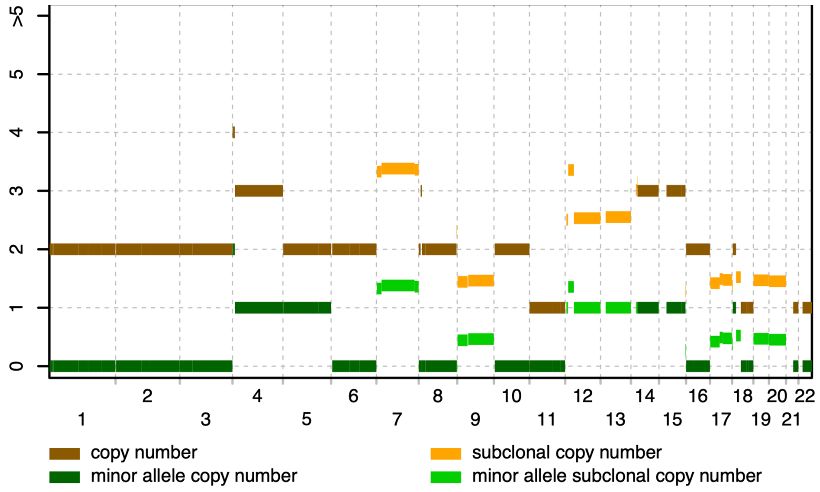

#### Supplementary Figures 02 A-F

#### Supplementary Figure 02 F – PanNET06

#### Metastases

M1

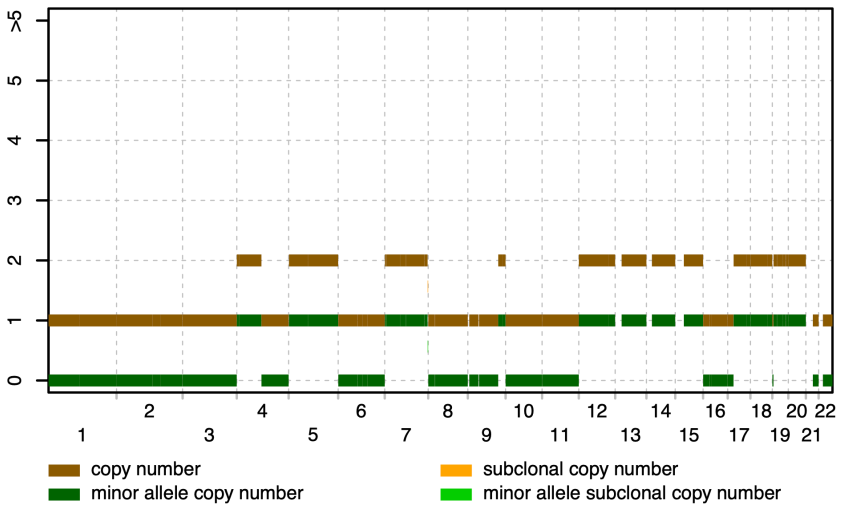


M2

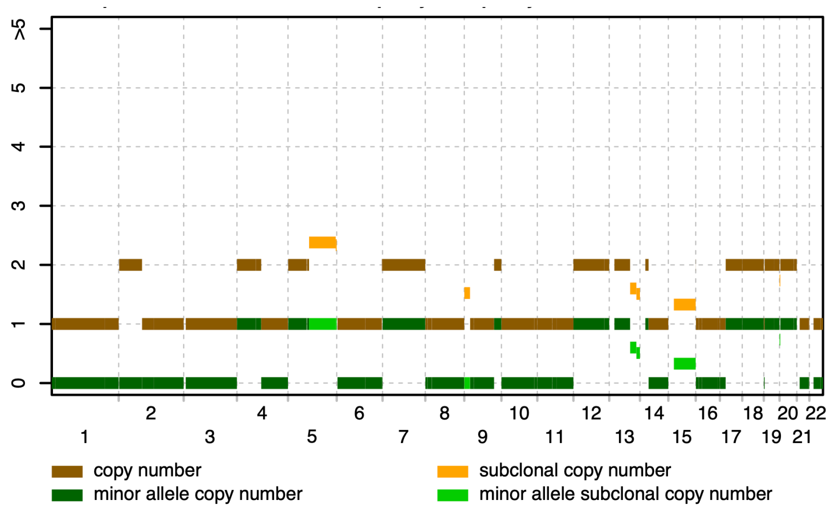

### Supplementary Figures 03 A-F

A

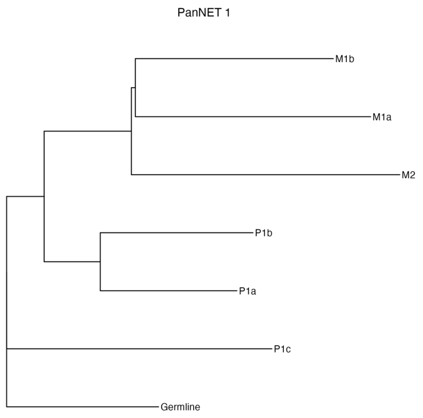


PanNET01: Metachronous metastases form outgroup, clearly descended from the same clone.

B

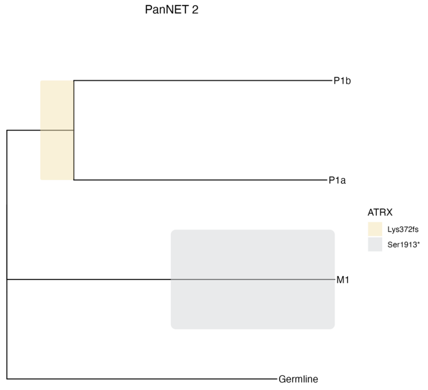


PanNET02: All samples share an inframe deletion in *MEN1* as well as a missense mutation in *TP53*. The two baseline samples share a frameshift mutation in *ATRX*, while the follow-up sample instead carries a distinct nonsense mutation indicating that the *ATRX* mutations occured later in disease course and exemplifying convergent/parallel evolution.

C

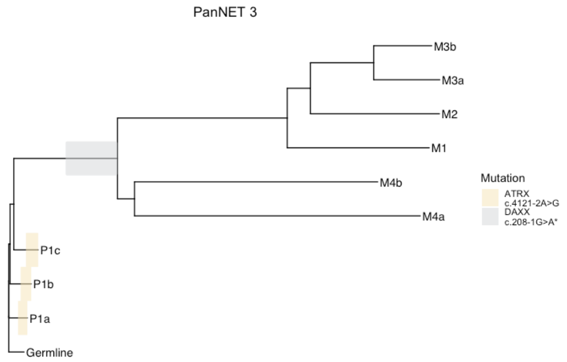


PanNET03: Extreme accumulation of mutations (increase of >100X) in metachronous/post-treatment/metastatic samples. The majority of these late mutations appear subclonal with low allelic fraction. Trunk length is comparable to that of other cases, but is insignificant compared to length of other branches in this tree. Risk of sample mixup can be excluded as all samples share many hundreds of mutations including a putative frameshift MEN1 driver mutation.

D

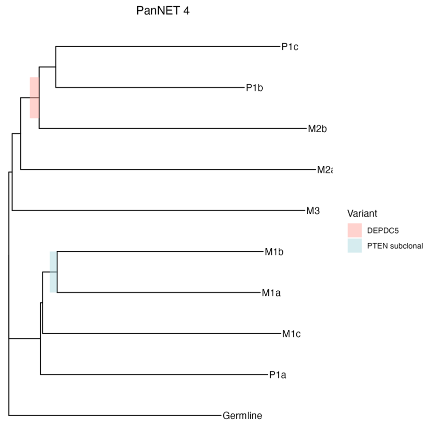


PanNET04: All: MEN1 nonsense, DAXX frameshift. M1a and M1b: PTEN nonsense. The SNV based phylogenetic analyses recapitulate the findings from DNA-methylation with highly similar tree topology indicating that the two processes occur in parallel rather than independently of each other.

### Supplementary Figures 03 A-F

**E**

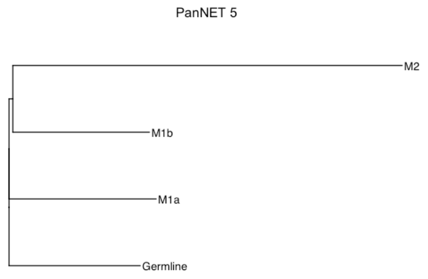


**F**

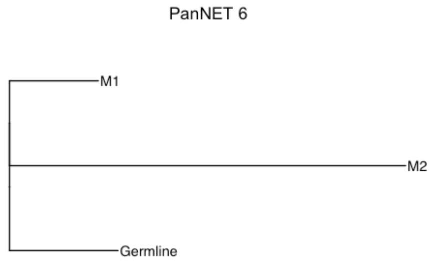


PanNET05: All samples share mutations in *MEN1*, *DAXX* and *TSC2*

PanNET06: The baseline sample M1 carries mutations in *TSC2* and *ARID1A* not found in the follow-up sample

### Supplementary Figures 04 A-H

**A**

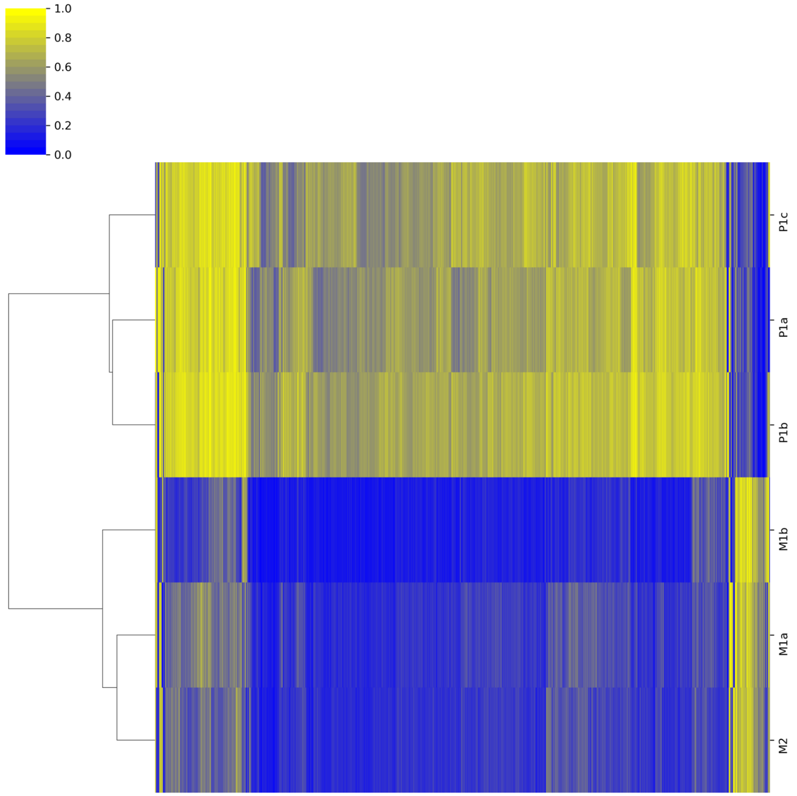


**B**

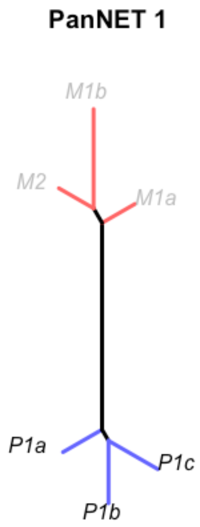

### Supplementary Figures 04 A-H

C

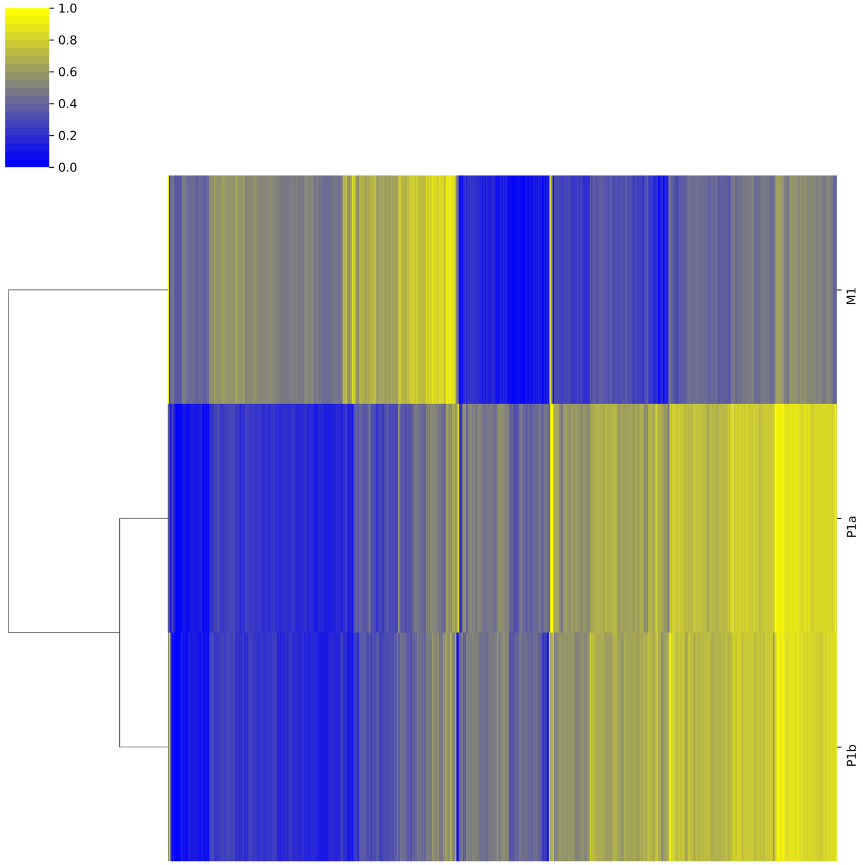


D

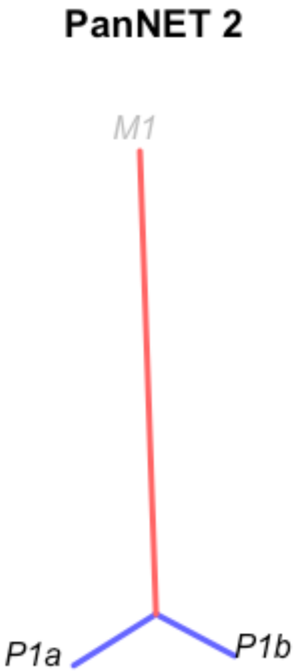

### Supplementary Figures 04 A-H

**E**

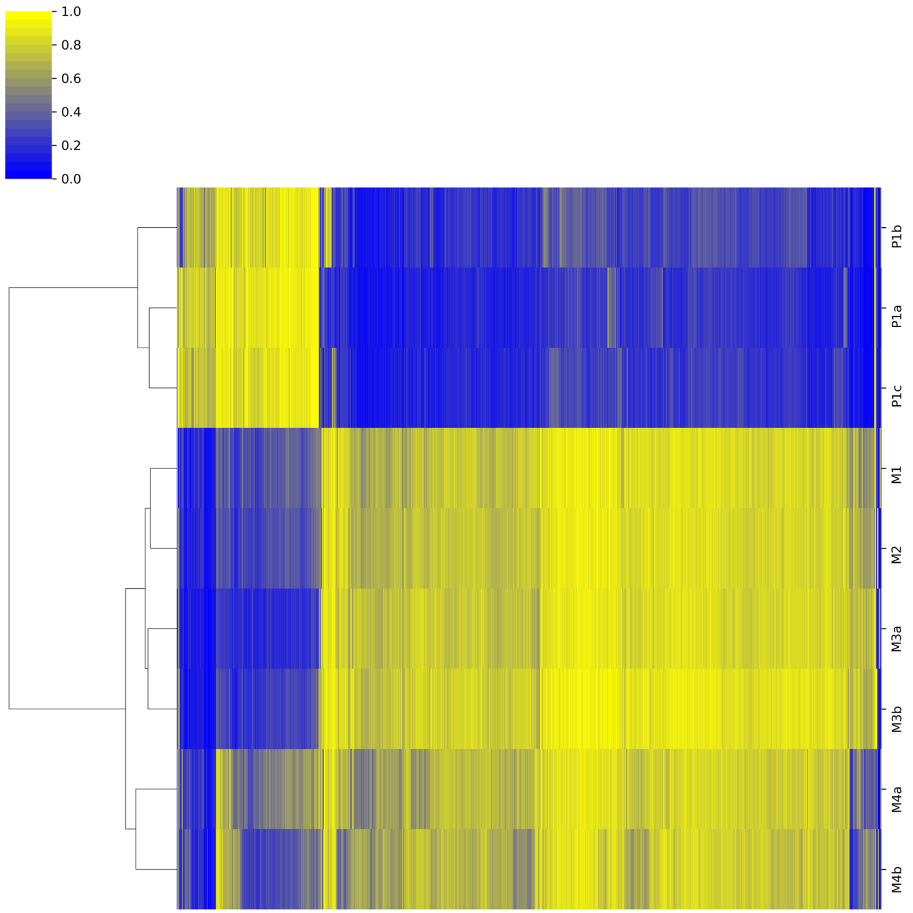


**F**

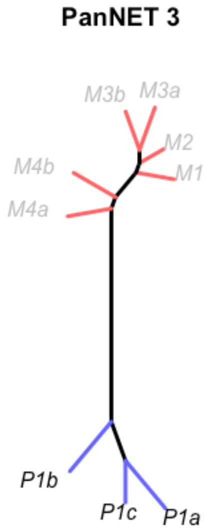

### Supplementary Figures 04 A-H

G

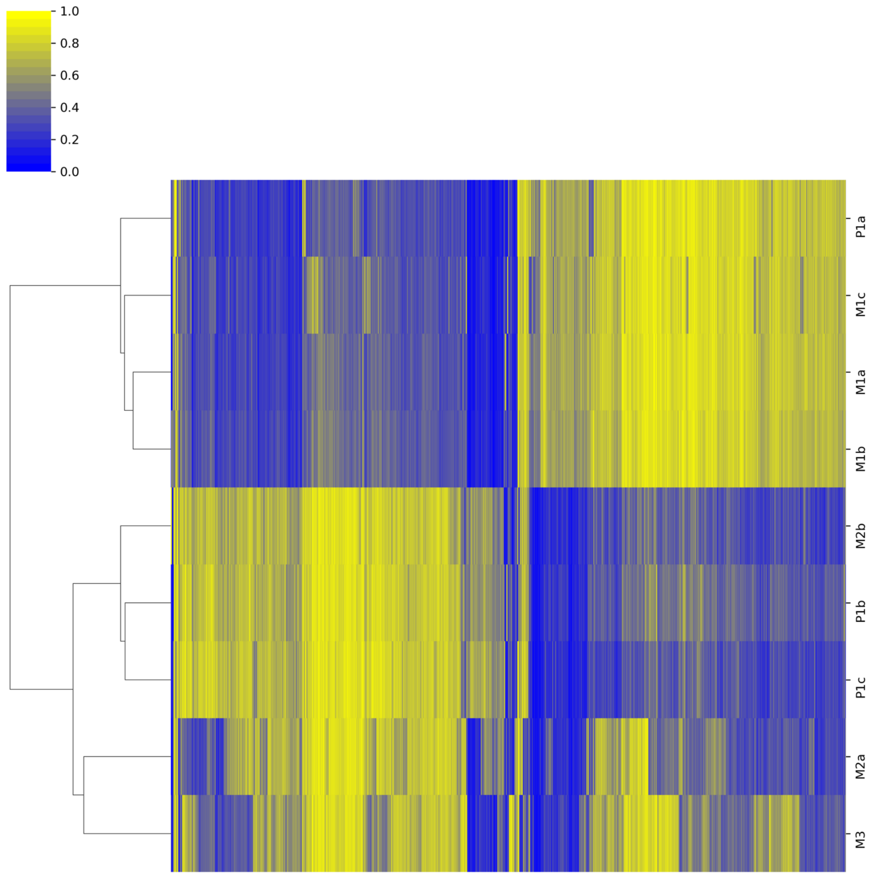


H

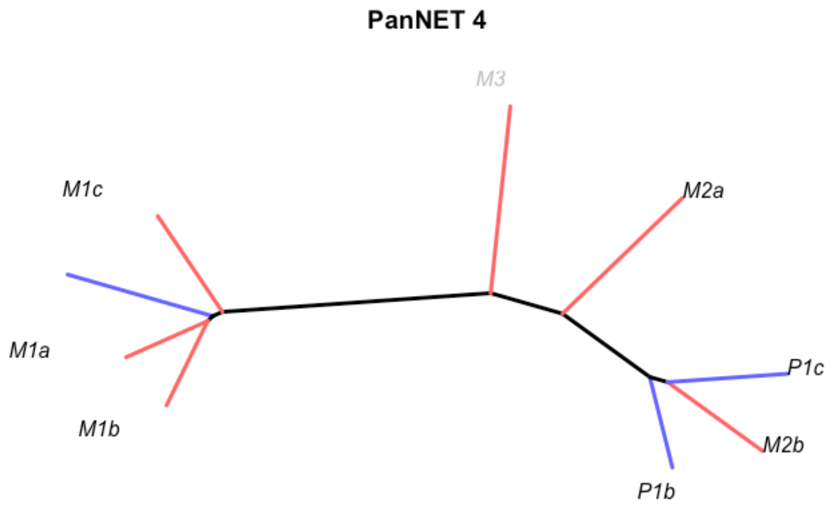

**Supplementary Figure 05**

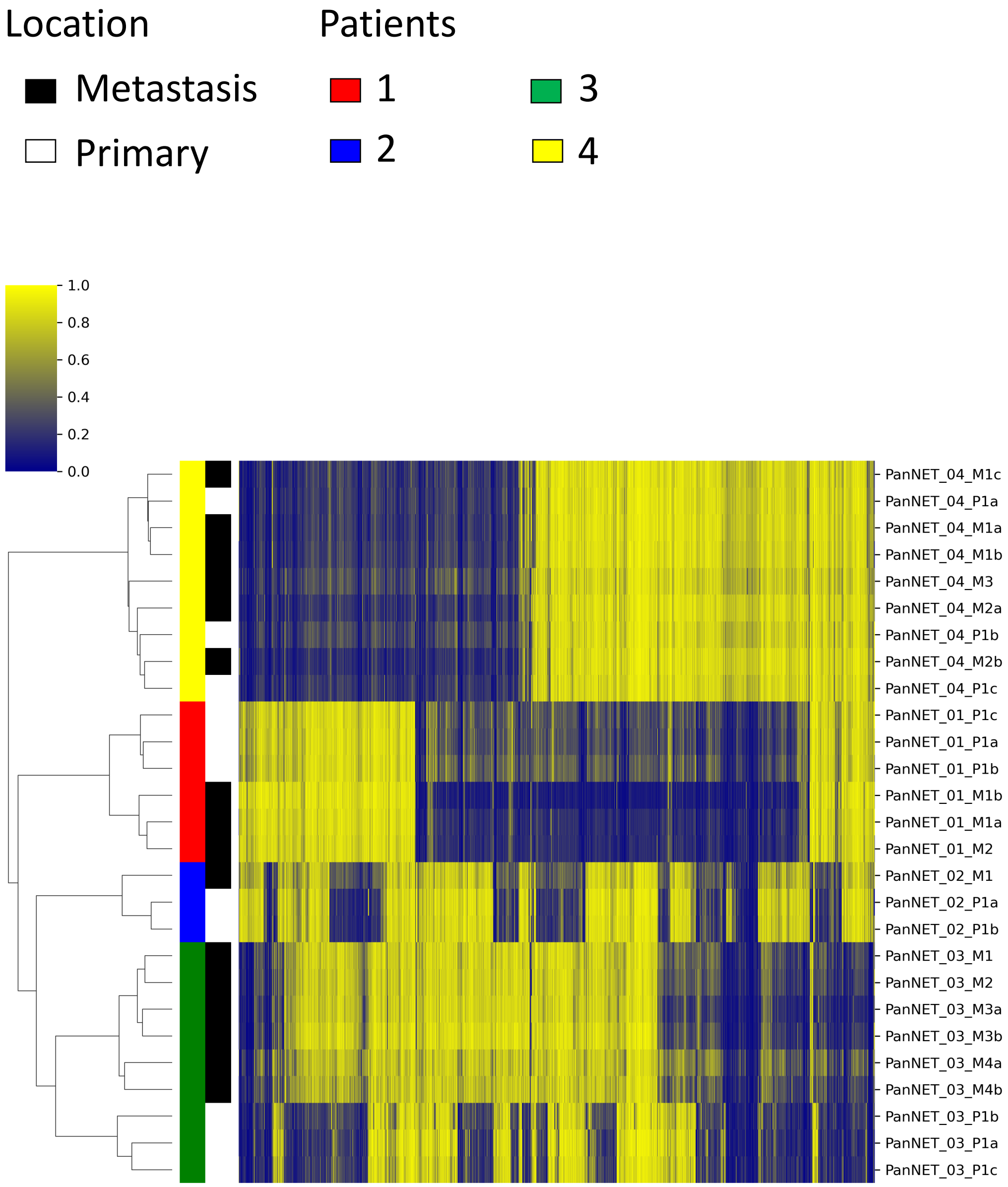

**Supplementary Figures 06 A-C**

### PanNET_03 RNA Seq

Patients

1 2 3 4

A

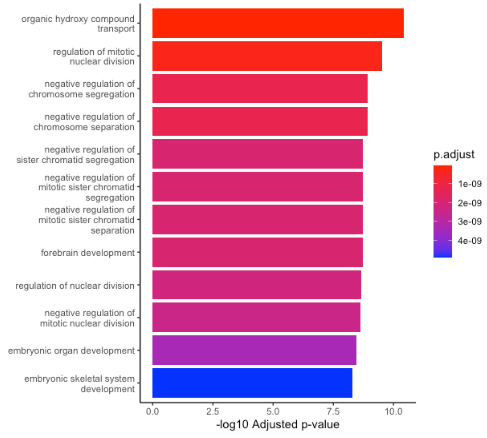


B

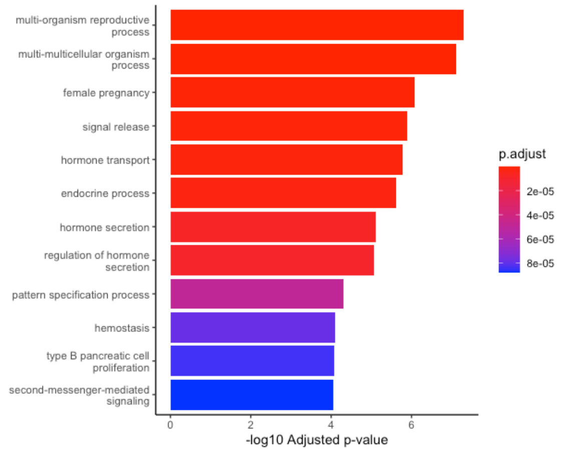


C

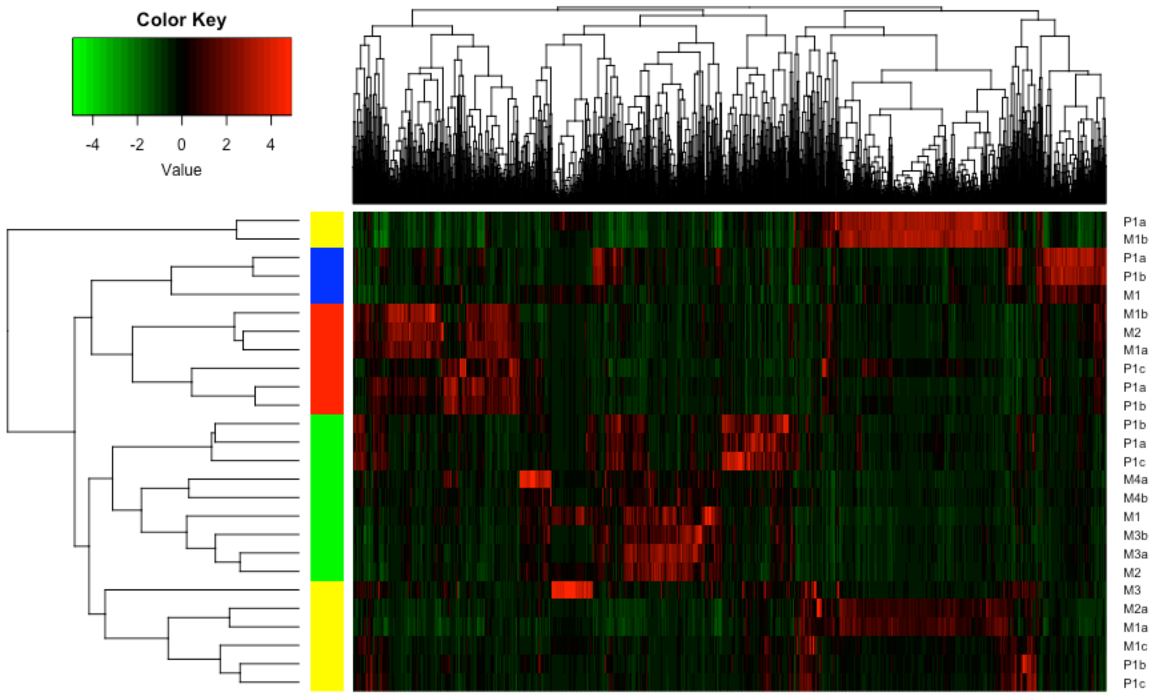

### Supplementary Figure 07 A-C

A

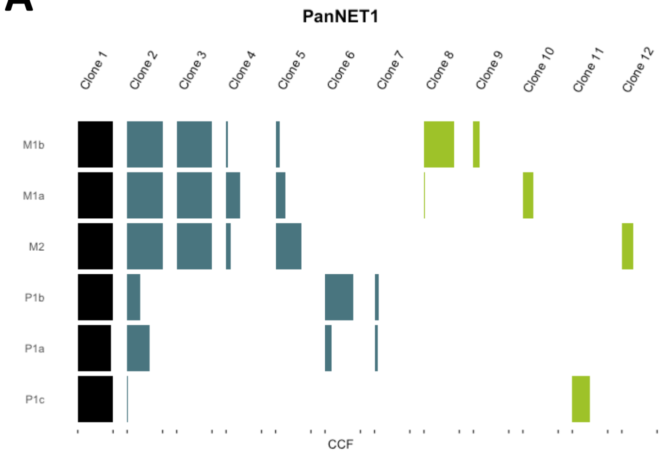


B

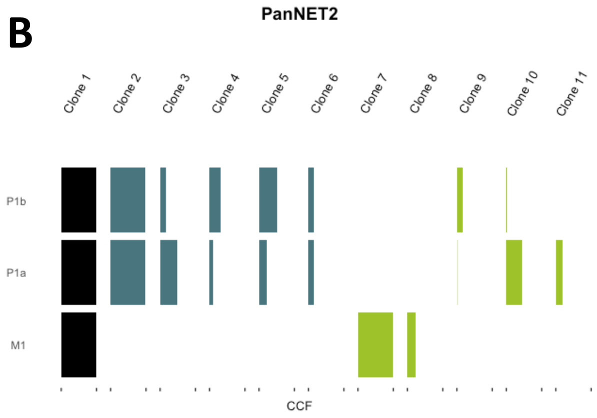


C

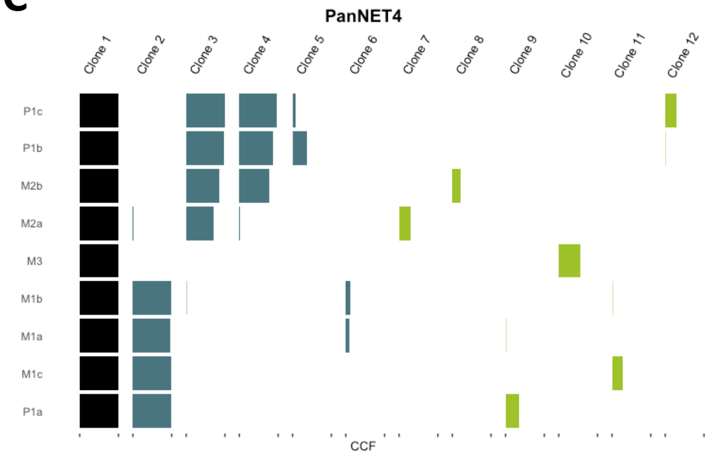

### Supplementary Figures 08A-I

A

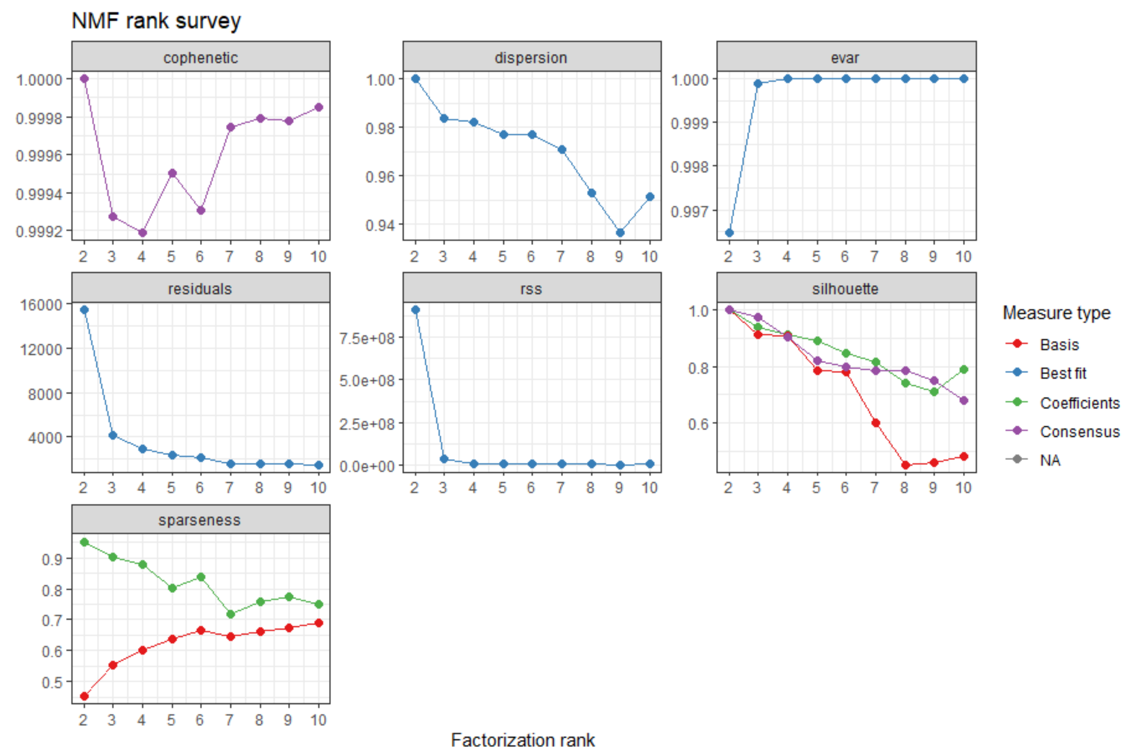


B

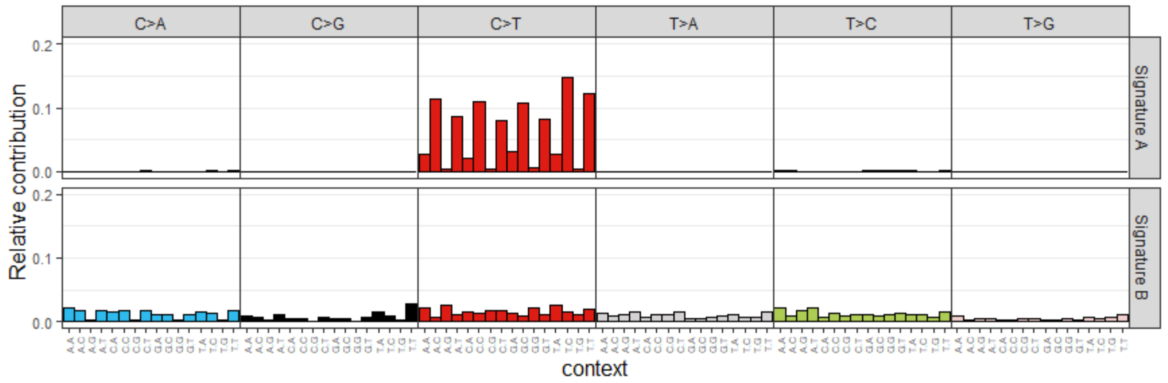


C

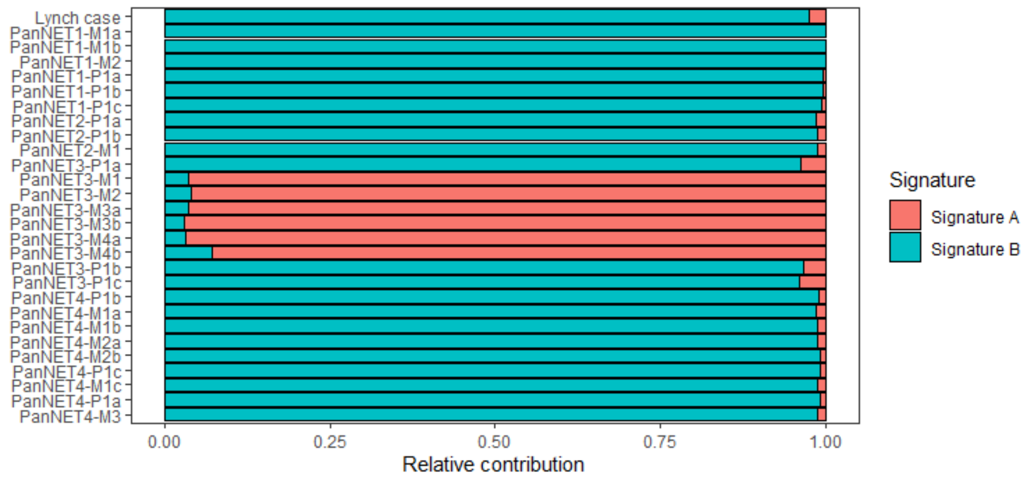

### Supplementary Figures 08A-I

D

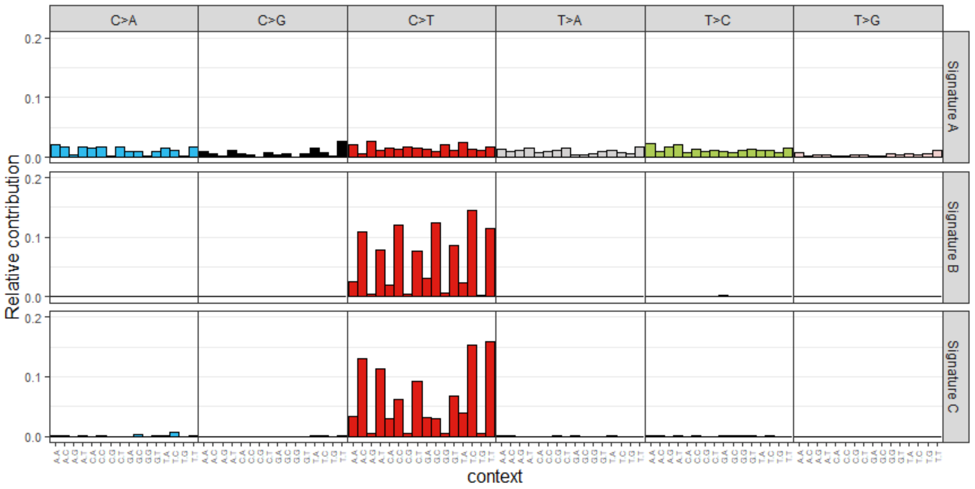


E

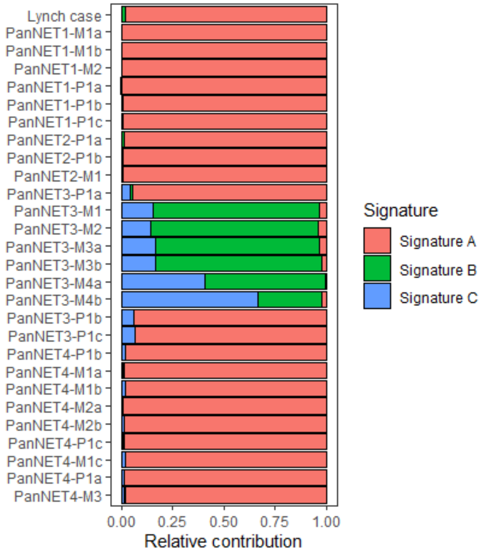


F

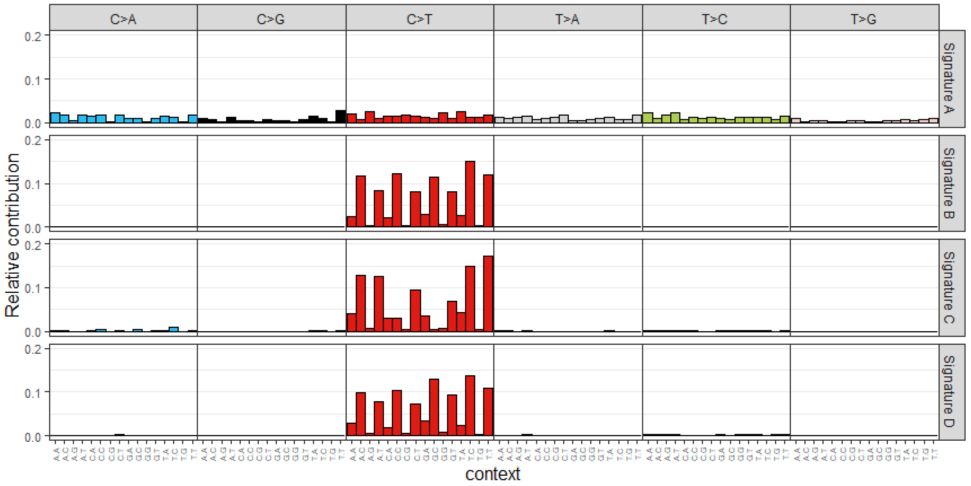


G

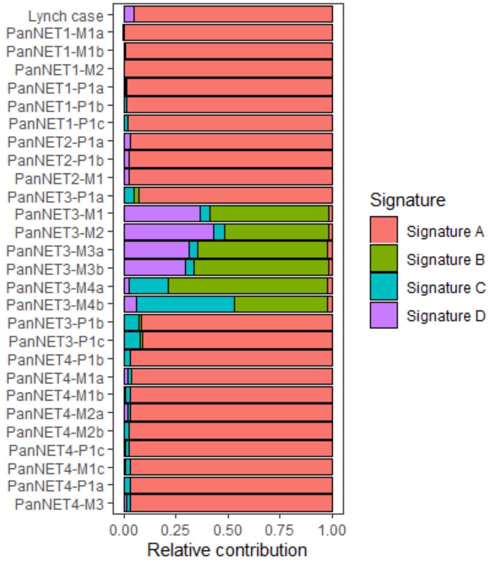

### Supplementary Figures 08A-I

## H

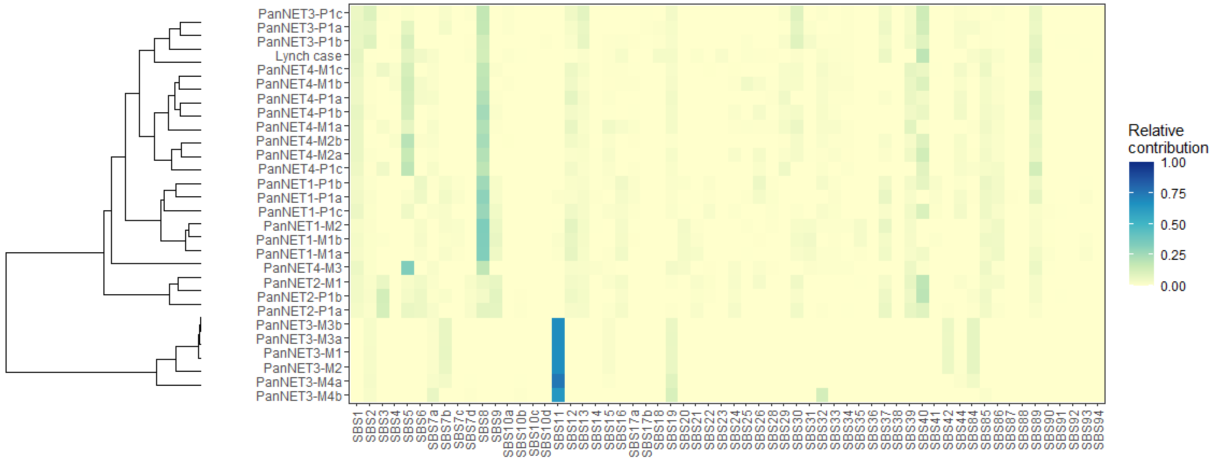


**COSMIC V3**

## I

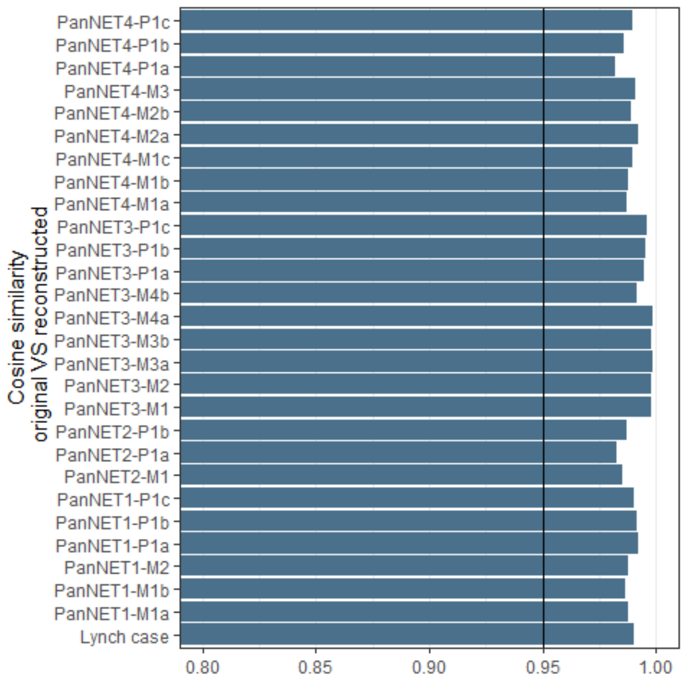

### Supplementary Figures 09A-B

## A

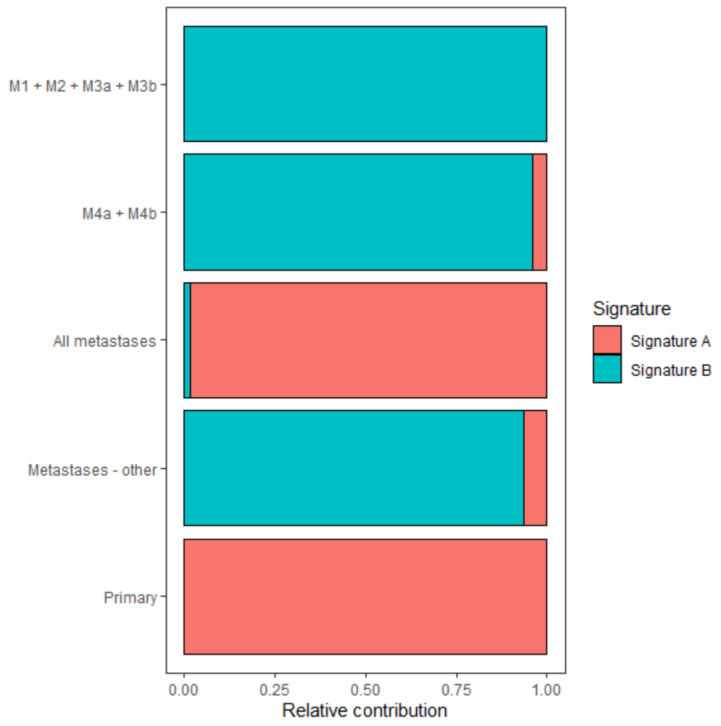


## B

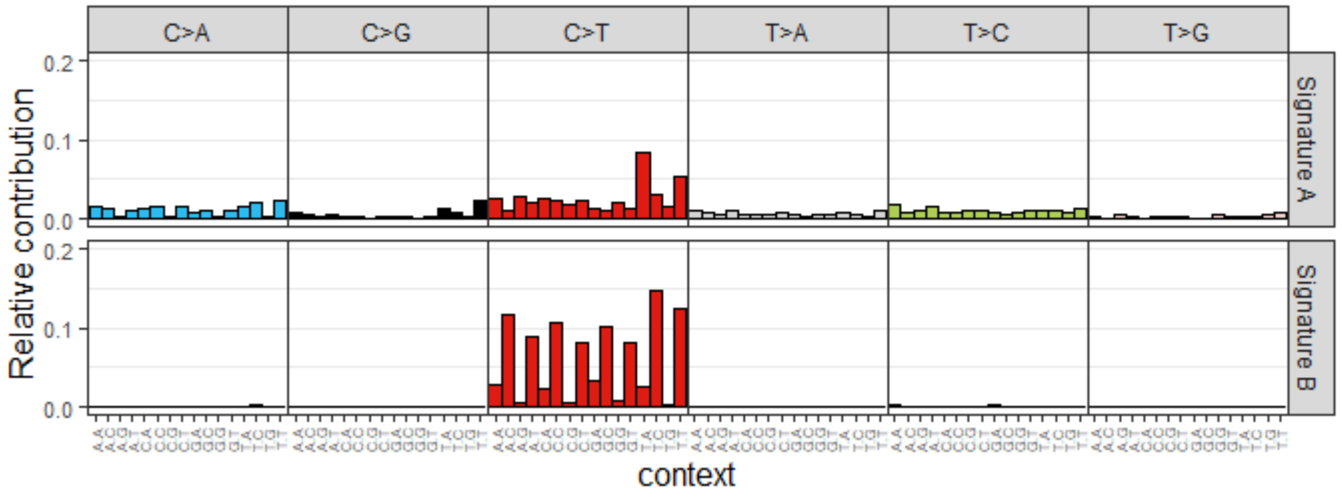

**Supplementary Figure 10**: DNA methylation difference between baseline and follow-up samples is greater in non-CpG-island sites

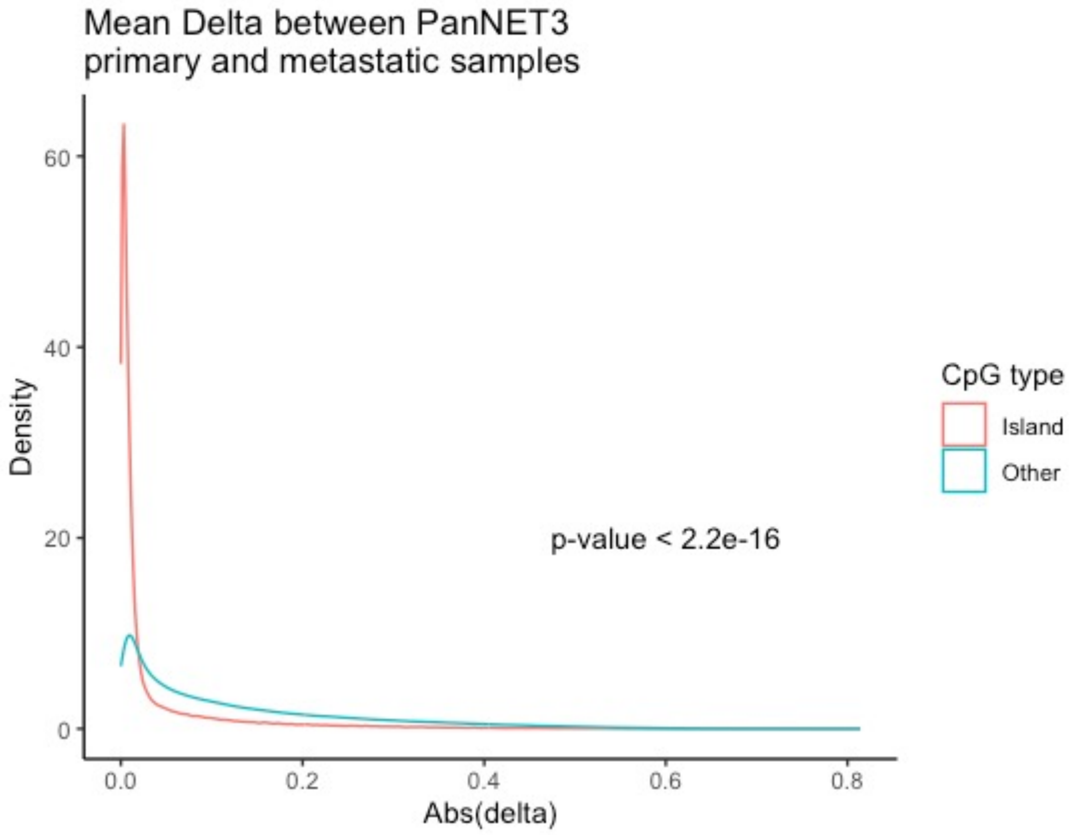

**Supplementary Figures 11A-B**

Pan-cancer clustering of Immune function enrichment in a cohort of 24 tumor types

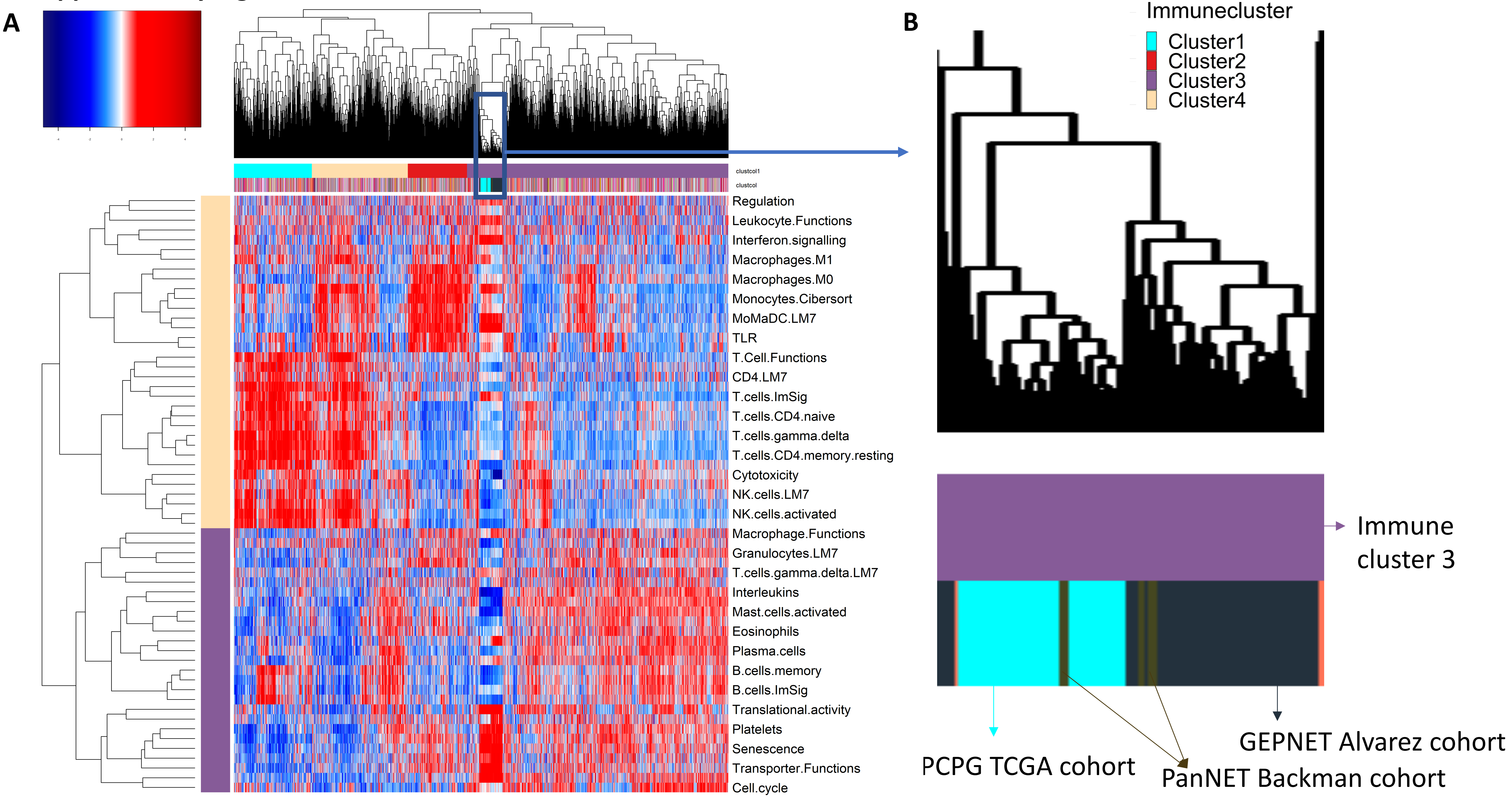

**Supplementary Figure 12**

Clustering of immune gene set enrichments (GSVA) across pancreatic tumor samples identified 3 clusters

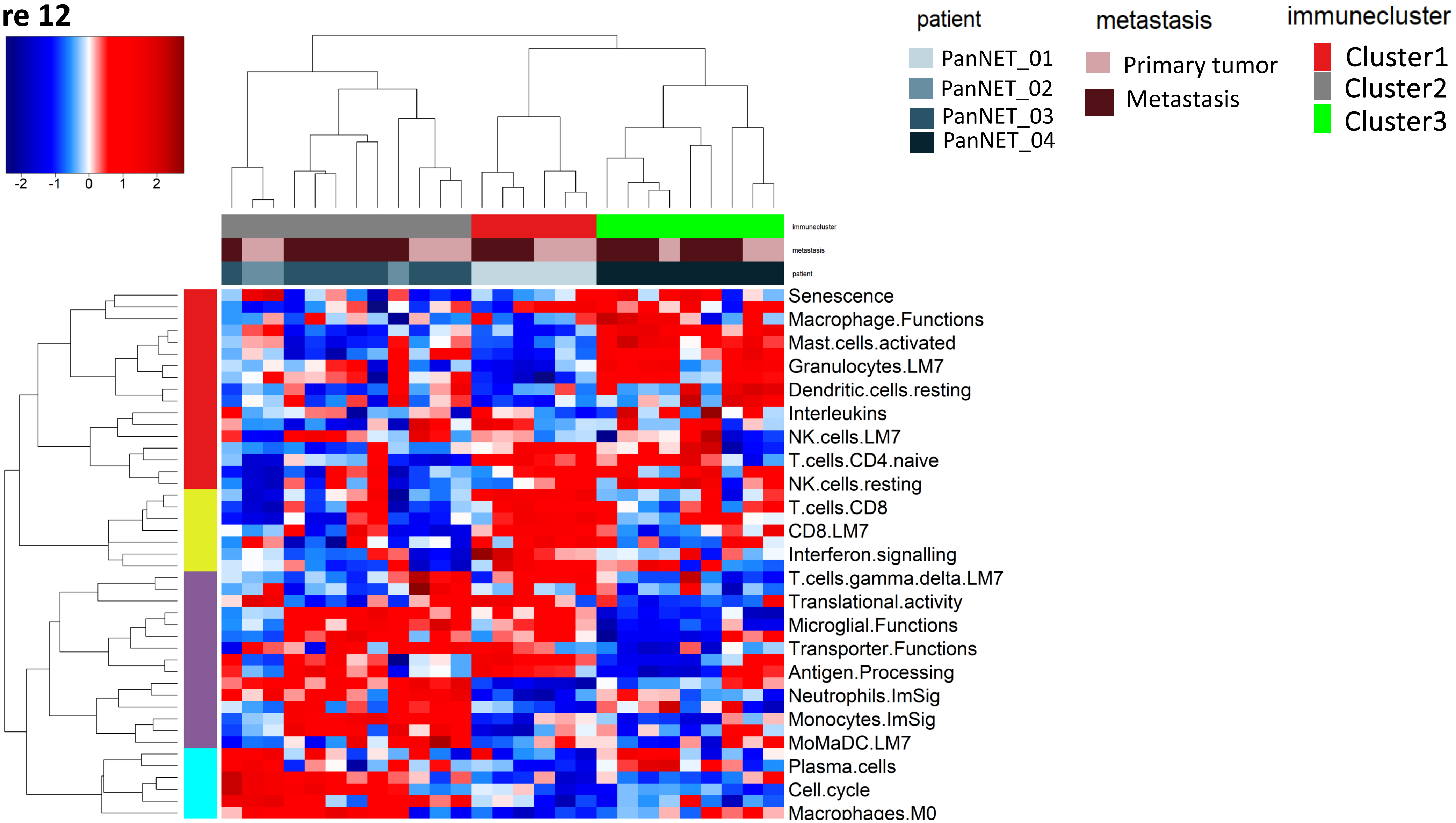

#### Supplementary Figures 13 A-D

**A** Pancreatic neuroendocrine tumors *n*=574

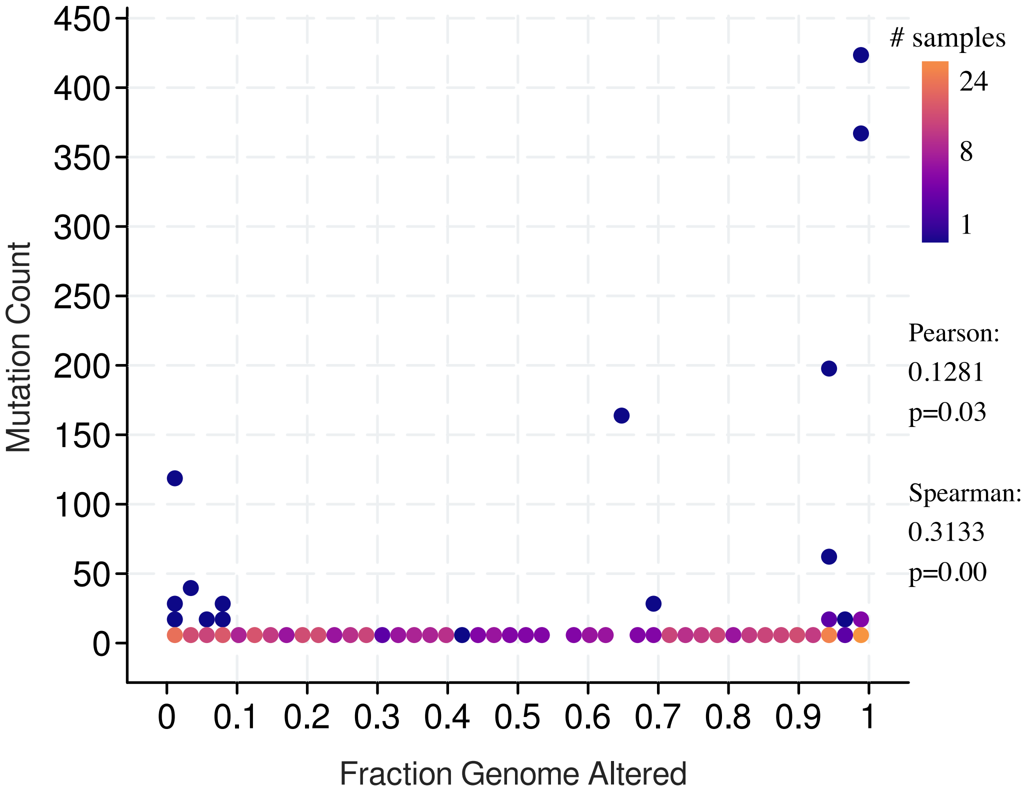


**B** Primary tumors *n*=278

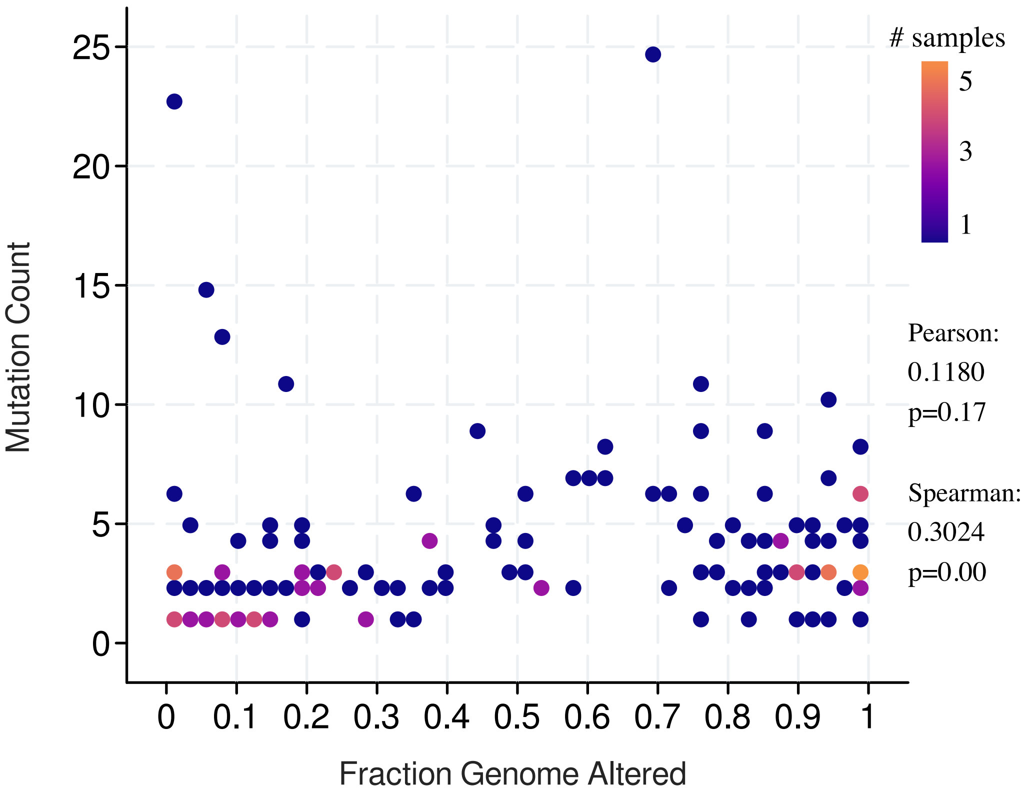

**Supplementary Figures 13 A-D**

**C** Metastases *n*=289

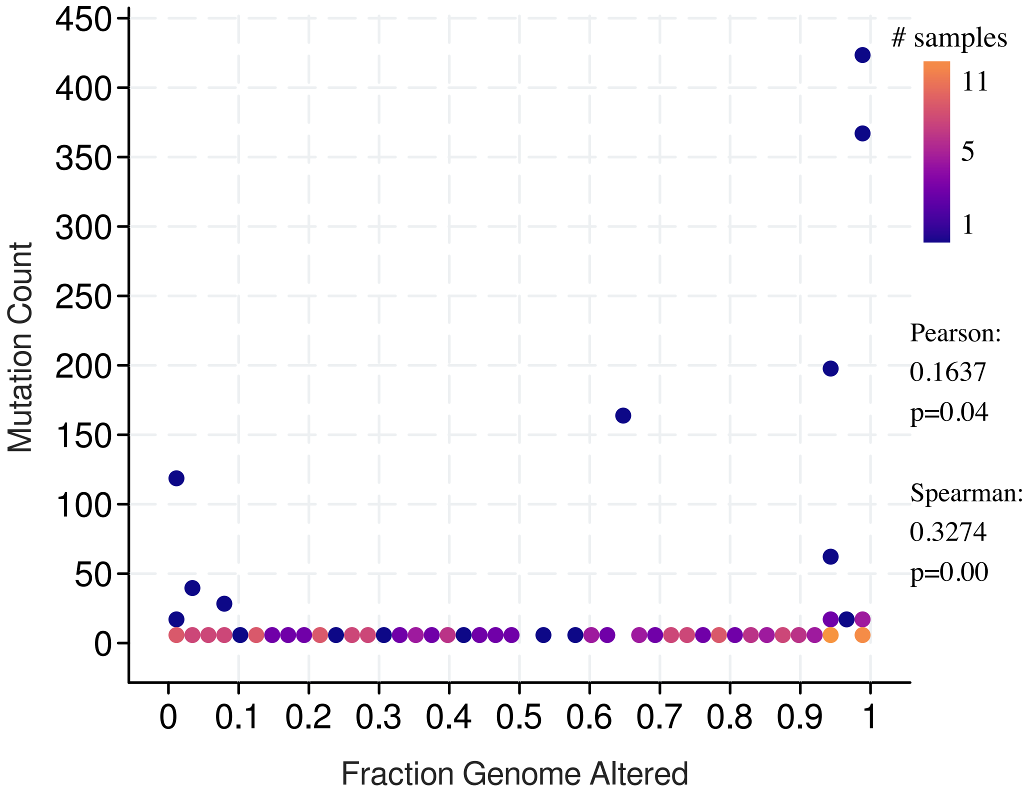


**D** *MSH6* mutated metastases n=8

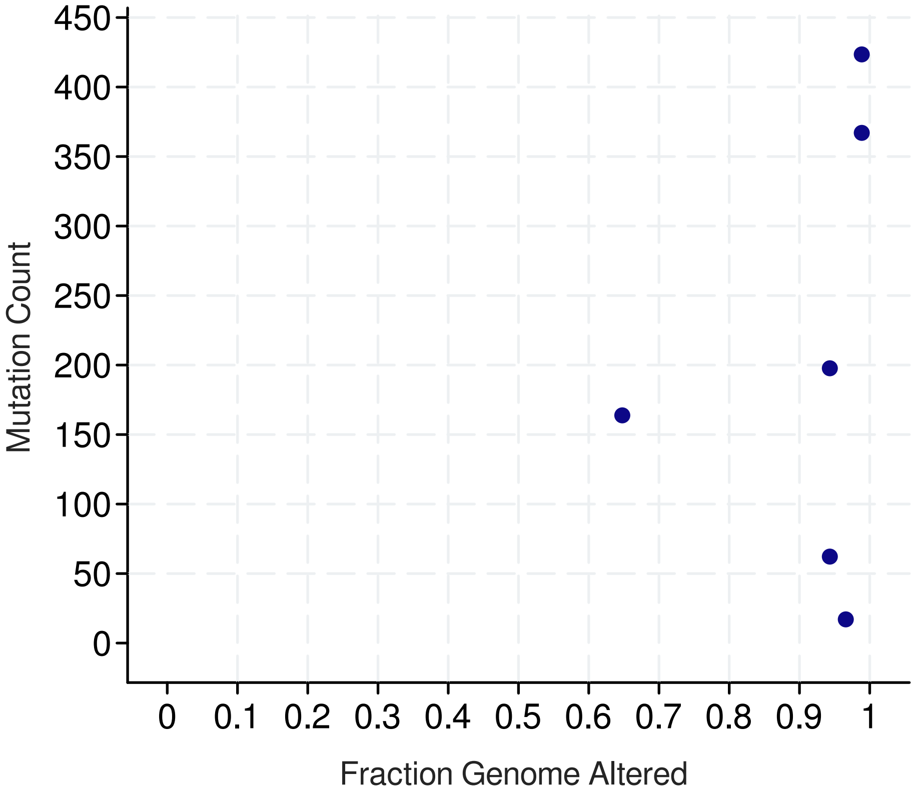

#### Supplementary Figure 14

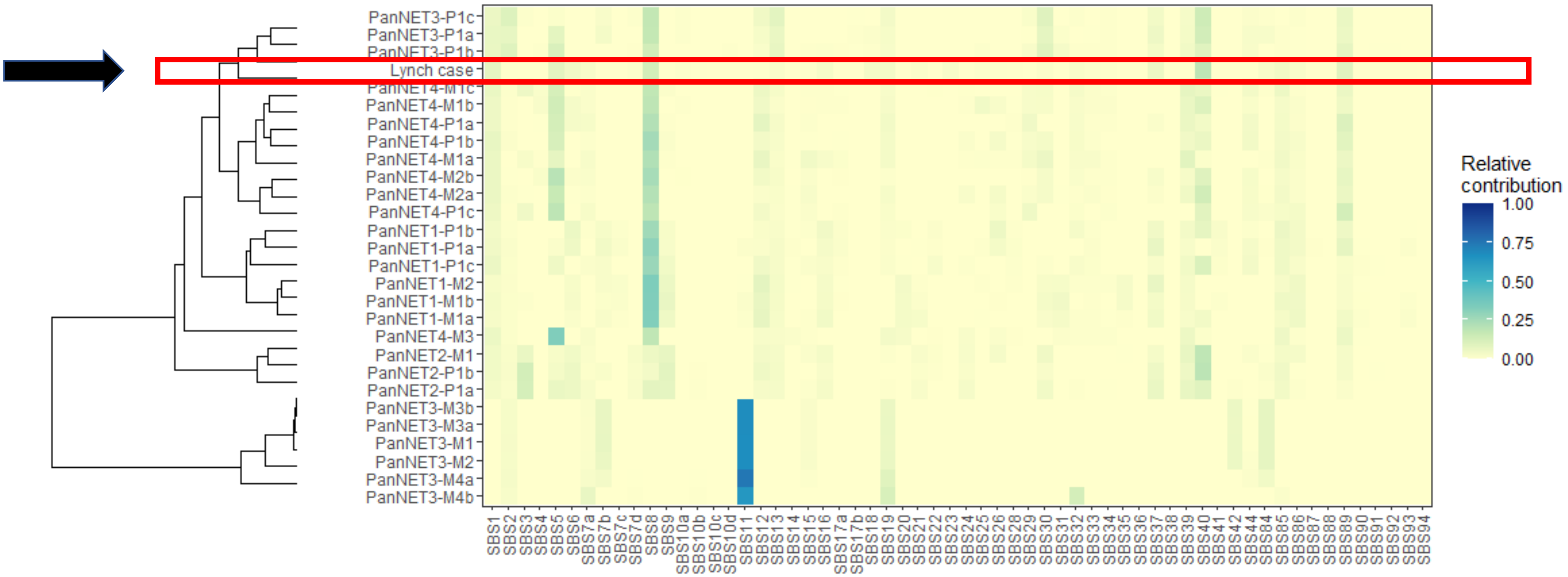

**Supplementary Figure 15 A-B** Major LOH events in PanNET3 and PanNET4 affect the same physical chromosome

**A** PanNET03

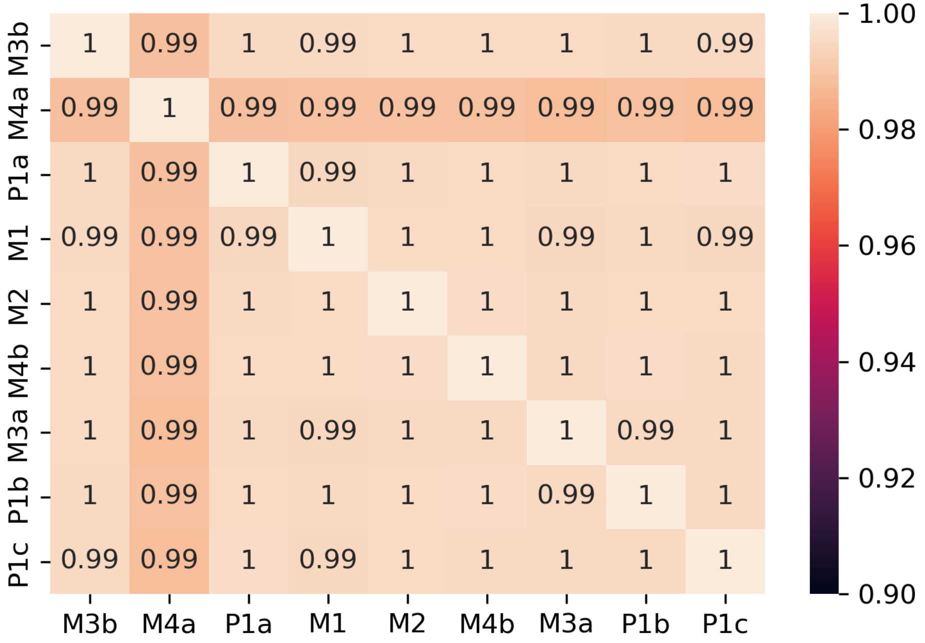


**B** PanNET04

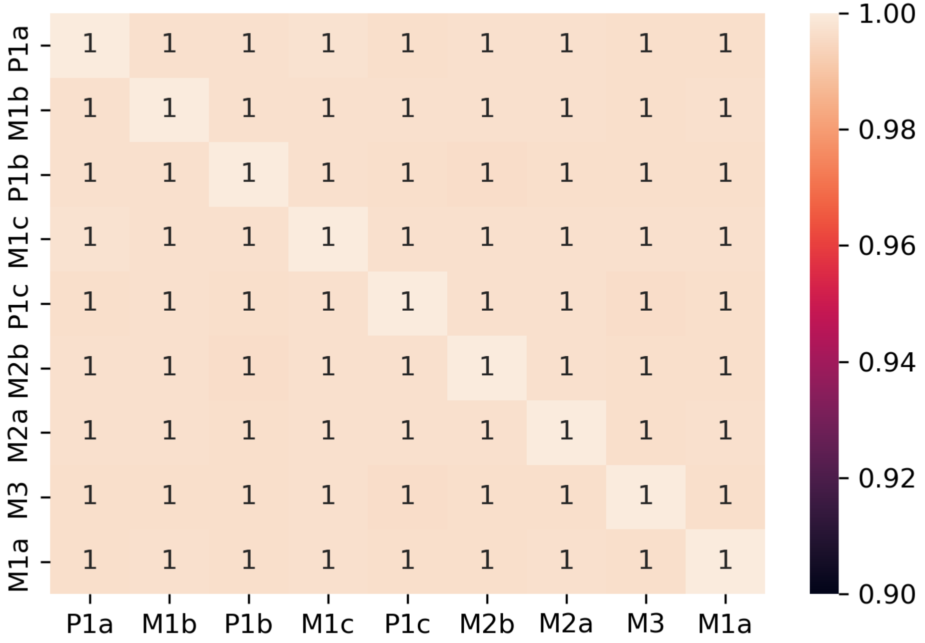
